# Transcutaneous Auricular Vagus Nerve Stimulation Reduces Inflammatory Biomarkers and May Improve Outcomes after Large Vessel Occlusion Strokes: Results of the Randomized Clinical Trial NUVISTA

**DOI:** 10.1101/2025.03.06.25323500

**Authors:** Osvaldo J. Laurido-Soto, Gansheng Tan, Susan Searles Nielsen, Anna L. Huguenard, Kara Donovan, Isabella Xu, James Giles, Rajat Dhar, Opeolu Adeoye, Jin-Moo Lee, Eric Leuthardt

## Abstract

**Background:** Inflammation plays a critical role in brain injury following acute ischemic stroke (AIS). Transcutaneous auricular vagus nerve stimulation (taVNS) has shown anti-inflammatory properties in various conditions, but its efficacy in AIS remains unexplored. We investigated if taVNS could mitigate post-AIS inflammation and its safety.

**Methods:** In this randomized, sham-controlled trial with blinded outcomes assessment, patients with anterior circulation large vessel occlusion (LVO) AIS were assigned to twice-daily taVNS or sham stimulation for five days or until discharge. Key inclusion criteria included age ≥18 years, National Institutes of Health Stroke Scale (NIHSS) ≥6, anterior circulation LVO, and enrollment within 36 hours of last known normal. Primary endpoints were changes in inflammatory biomarkers (white blood cells and cytokines including interleukins (IL)-1β, 6, 10, 17α, and tumor necrosis factor alpha (TNFα) measured at baseline and Days 1, 3, 5, and 7, and taVNS safety. Secondary exploratory endpoints included change in NIHSS, 90 day modified Rankin score (mRS), and safety (bradycardia, hypotension, infection, and death).

**Results:** Thirty-five patients (17 taVNS, 18 sham) were enrolled. The taVNS group showed a significant rate of change in normalized aggregate pro-inflammatory cytokines and interleukin-6 levels compared to sham (p=0.04 and p<0.001, respectively). Each 1 pg/mL reduction in interleukin-6 correlated with a 0.798-point improvement in NIHSS in the taVNS group (95% confidence interval [0.077, 1.518], p = 0.031]), with no significant correlation in the sham group. IL-1β, 10, 17α, and TNFα showed reduction in cytokine levels, but did not reach statistical significance. There were no statistically significant differences amongst mRS and safety outcomes between both groups.

**Conclusions:** taVNS safely reduced post-AIS inflammation in anterior circulation LVO stroke patients, demonstrating biological effects. Secondary analyses also found potential effects in NIHSS improvements. These promising findings warrant further investigation in larger trials.

**ClinicalTrials.gov ID:** NCT05390580, https://clinicaltrials.gov/study/NCT05390580

## Introduction

Acute ischemic stroke (AIS) due to large vessel occlusions (LVOs) lead to disproportionate morbidity and mortality without emergency treatment.^1^ Unfortunately, even after successful recanalization with mechanical thrombectomy, i.e., reperfusion of the primary vessel occluded [Thrombolysis In Cerebral Infarction score (TICI) 2B, 2C, or 3],^2^ infarcts can continue to increase in size or develop hemorrhagic transformation,^3,4^ leading to poor outcomes, highlighting the need for adjunct therapies to minimize ischemia progression and morbidity for both recanalized and non-recanalized patients. Neuroinflammation has been recognized as an important contributor to ischemic brain injury.^5^ In the acute phase of AIS, inflammatory biomarkers elevation, such as pro-inflammatory cytokines [e.g., interleukin (IL) 6 (IL-6)],^6–10^ are associated with exacerbating brain injury. Although ongoing trials are occurring in AIS, most immunomodulatory drug trials so far have been disappointing due to limited efficacy and/or medication side effects.^11–14^ Thus, finding adjunct treatments that are easy to administer and safe is of the utmost importance.

Vagal nerve stimulation (VNS) has been established to have anti-inflammatory effects in several inflammatory conditions in animal models, including aneurysmal subarachnoid hemorrhage and inflammatory bowel disease.^15–18^ In animal models of cerebral ischemia and reperfusion injury, VNS not only reduced infarct size, but also improved neurological outcomes.^19–21^ In practice, most FDA-approved VNS are currently administered through surgical implantation of an electrode placed directly on the vagus nerve in the carotid sheath in the neck. While appropriate for chronic conditions such as depression, epilepsy, and chronic stroke recovery,^22^ implantable solutions are impractical in the emergent setting of AIS. In this context, noninvasive transcutaneous auricular VNS (taVNS) offers a possible solution given its immunological effects and extremely low morbidity.^23,24^ Based on encouraging animal studies in AIS, demonstrated safety of taVNS in the critical care settings,^25^ and evidence of efficacy in subarachnoid hemorrhage,^26^ we conducted the “Neuromodulation Using Vagus Nerve Stimulation Following Ischemic Stroke as Therapeutic Adjunct” (NUVISTA) trial. This prospective randomized sham-controlled with blinded outcomes clinical study aimed to quantify the effect of taVNS on plasma inflammatory biomarkers in patients with LVO and explore its impact on post-stroke patient outcomes.

## Methods

### Study population

All patients with incident AIS due to LVO who were admitted to Barnes Jewish Hospital in St. Louis, Missouri, between November 2022 and May 2024 were screened for eligibility. Inclusion criteria included: age≥18 years, acute anterior circulation LVO [Internal Carotid Artery (ICA), Middle Cerebral Artery first branch (M1), Middle Cerebral Artery second branch (M2)], National Institutes of Health stroke scale (NIHSS) score ≥6, pre-morbid modified Rankin score (mRS) ≤2, randomization <36 hours (hrs) from symptom discovery/last known normal (LKN), expected life expectancy >3 months, and no active cancer or immunosuppressive/modulating therapy, concomitant infections or inflammatory states (i.e. chronic autoimmune diseases), hypotension, or bradycardia on arrival. The patient/family, the medical team, and the outcomes assessor were blinded to which arm the patient was enrolled (**Figure 1).** Full *<u>Inclusion</u>* and *<u>Exclusion</u>* criteria shown in **Supplementary Table 1.**

**Figure 1.**
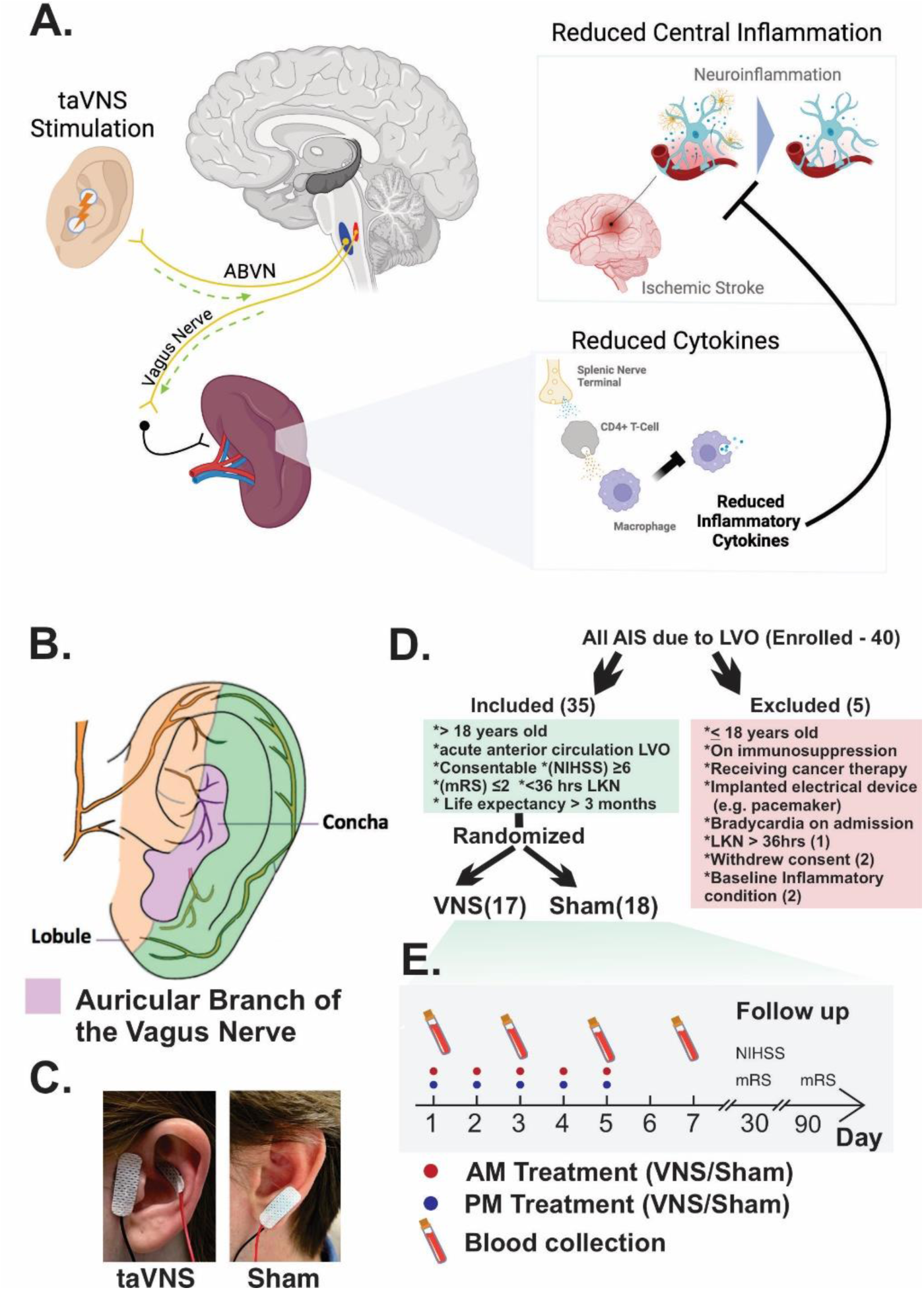
Rationale and trial protocol. **A**. The proposed mechanism of taVNS in acute ischemic stroke patients. Electrical stimulation of the auricular branch of the vagus nerve activates the cholinergic anti-inflammatory pathway. When the dorsal vagal complex receives input from the auricular branch of the vagus nerve, it activates efferent vagal fibers that project to the spleen. This process suppresses the production and release of pro-inflammatory cytokines like TNF-alpha, IL-1beta, and IL-6 and reduces central inflammation. **B**. Distribution of the auricular branch of the vagus nerve on the outer ear.^1^ **C**. Electrode placement for taVNS stimulation and sham group stimulation. **D**. Inclusion and exclusion criteria. **E**. Timeline for treatment and measurement of primary outcomes.

### Protocol

Patients were randomized to taVNS stimulation for 20 minutes every 12 hours vs sham electrical stimulation according to the same schedule. Our basic stimulation parameters were: intensity – 0.5 mA, frequency – 25 Hz, and pulse width −200 μs (**Figure 1)**.^27^ Patients continued left ear taVNS or left ear sham stimulation for five days (10 total stimulations), discharge, or death, whichever came first, and received standard procedural and medical care. Full protocol and device information shown in **Supplement Information 1** and CONSORT checklist^28^ adherence as per **Supplement Information 2**.

### Assessment of patient characteristics and standard treatment

Baseline patient characteristics were documented, including age, sex, race, smoking history, comorbidities, admission baseline NIHSS, and pre-stroke modified Rankin Scale (mRS) score. Each stroke event was characterized by laterality, occlusion type, and acute interventions. Treatment documentation included thrombolytic administration and thrombectomy details. In thrombectomy cases, reperfusion outcomes were assessed using the TICI score at the end of thrombectomy.^2^ This score was analyzed both as a semi-continuous measure of reperfusion percentage and as a dichotomized outcome (successful reperfusion defined as TICI ≥2B).^2^

### Brain imaging and assessment of biomarkers of cerebral edema and infarct size as a baseline variable

Head CT scans were obtained at baseline (within 12 hrs of last known normal) and follow-up, including at 24 +/-12 hrs post-intervention. Baseline scans were available for 13 (76.5%) treatment and 13 (72.2%) sham patients. All patients had at least one follow-up scan. Following established protocols,^29,30^ we analyzed the scans for measures of cerebral edema and infarct size, including cerebral spinal fluid volume ratio and relative hemispheric volume to utilize as baseline measures. We also assessed for hemorrhagic transformation and malignant edema.

### Assessment of blood biomarkers of inflammation

Assessment of blood biomarkers of inflammation was the primary outcome. Total blood levels (pg/ml) of five cytokines were assessed: four ILs, specifically IL-1β, IL-6, IL-10, and IL-17α, and tumor necrosis factor alpha (TNFα). White Blood Cell (WBC) counts (K/cumm) were also measured. The full blood draw protocol can be found in **Supplement Information 3**. All five cytokines were measured at baseline (Day zero/<24 hrs) and every other day for the week thereafter (Day 1, Day 3, Day 5, and Day 7) or until hospital discharge. Protein concentrations were reported as pg/mL. As a summary measure, the mean of the z-score for each of the five cytokines (hereafter “cytokine score”) was calculated. Z-scores were used to give similar weight to each cytokine, as IL-6 pg/ml levels were much greater than the other cytokines and therefore dominated a summary measure based on absolute values. All five cytokines were included together, rather than excluding IL-10 due to its anti-inflammatory characteristics, based on preliminary analyses that indicated this cytokine generally rises similarly to the other cytokines;^31^ and we also analyzed pro-inflammatory cytokines in isolation. Cytokine quantification methods can be found in **Supplement Information 4**.^32^

### Assessment of potential adverse effects

Due to the potential cardiovascular effects of VNS,^33^ safety outcomes were monitored, including hypotension (systolic blood pressure <80 mmHg) and bradycardia (heart rate <50 beats per minute). Vital signs were recorded at multiple times around each stimulation session: 5 minutes before, every 5 minutes during, and 5 minutes after stimulation. Additionally, routine vital sign monitoring was performed every 12 hours throughout the hospital stay.

### Assessment of neurological outcomes

The NIHSS total score and subscores were assessed at presentation/admission and daily thereafter until discharge or death, as well as on Day 30 +/- 7 days from LKN. Clinical assessments were performed by NIHSS certified stroke coordinators, nurses, and providers and documented in the medical record. The mRS was evaluated on Days 30 and 90 +/- 15 days from LKN over the phone, utilizing the Joint Commission National Quality Measures.^34^ Additional clinical outcomes included hospital length of stay, while discharge disposition was categorized as either favorable (home, home with health services, or inpatient rehabilitation) or unfavorable (skilled nursing facility or long-term acute care hospital), or death before discharge. Patients were followed for 105 days after admission or until death,^34^ with readmissions to the hospital system and deaths at any location documented during this period. All vital status information and death dates were verified through electronic medical records.

### Statistical analysis

Statistical analyses were performed using Stata/MP version 18 and Python packages Scipy and Statsmodels. All continuous variables were retained as continuous, except where noted. Multivariable regression was used to examine treatment group associations with outcomes, except for rare secondary outcomes/adverse effects. Following FDA recommendations,^35^ an intention-to-treat (ITT) approach was implemented. Primary ITT analyses were conducted without transformation of continuous outcome variables because ≥35 observations were available.

### Statistical analysis of rate of change in WBC, cytokines, and neurological outcomes in taVNS vs. sham treatment

Longitudinal data were analyzed using mixed effects models per the FDA,^36^ with person as a random effect, given multiple records per person. For cytokine models, assay plate was included as an additional random effect. This approach accommodated irregularly-timed points, less restrictive missing data assumptions, and time-varying variables. To complement our primary ITT approach, treatment also was handled as time-varying when noted, with assignment occurring at randomization. All longitudinal models included treatment, time since LKN, and their interaction term(s) as primary independent variables to test whether the pattern of change over time differed between the taVNS and sham groups. Time was modeled quadratically for WBC and cytokine outcomes to capture the known U-shaped associations with time^37^ when modeling these outcomes and linearly for in-hospital NIHSS scores based on panel data plots. For NIHSS a single model was constructed to estimate the rate of change (change in NIHSS per day) in each of the two treatment groups and to formally test whether these rates were different. For 30-day NIHSS changes (vs. baseline) and 90-day mRS changes, we used the parallel model, with actual assessment days scaled to standardize the evaluation period to 30 or 90 days, respectively. We characterized the daily change in each WBC or cytokine outcome using a similar model expanded to include the quadratic terms. Treatment-time interactions were tested using likelihood ratio tests comparing models with and without interaction term (NIHSS, mRS) or terms (WBC, cytokines), with significance at two-sided α=0.05. These analyses were adjusted for baseline covariates,^38^ including baseline NIHSS (except in NIHSS models, which included this observation), time from LKN to thrombus removal time or recanalization attempt (groin puncture time), and percent reperfusion (0-100%) based on TICI score^2^ to obtain a single semi-continuous measure. In sensitivity analyses, we also performed the analysis with a dichotomized TICI scale (reperfused ≥2B, not reperfused <2B) or restricted data to the first five days post-LKN (when most stimulations and hospitalizations had ended). Finally, we explored whether any difference in longitudinal patterns between taVNS and sham depended on laterality, occlusion type, or any of the *a priori* adjustment variables, i.e., whether any time-treatment interaction depended on one of these third variables. Laterality was of particular interest due to previous research suggesting differential VNS effects based on the side of stimulation^39,40^ and its influence on NIHSS subscore patterns. Full model descriptions can be found in **Supplement Information 4**.

### Statistical analysis of difference in cytokine levels between taVNS and sham groups

We conducted post-hoc t-tests to compare cytokine levels between groups on each day. We chose to structure further analysis utilizing IL-6 based on prior literature suggesting IL-6 is a key cytokine associated with stroke outcomes and prior reduction of IL-6 with taVNS in aneurysmal subarachnoid patients.^26,41^

### Statistical analysis of the association between change in cytokines and change in clinical outcomes

We investigated the association between IL-6 levels and clinical outcomes, or their respective changes, to understand the short- and long-term functional implications of reducing inflammatory cytokines. We first verified if the reduced pro-inflammatory cytokine was associated with improving neurological outcomes using regression with changes in NIHSS scores as the dependent variable and changes in IL-6 as the independent variable. In this analysis, we paired Day 3 and Day 5 NIHSS and IL-6 measurements based on the measured time and then calculated the change by subtracting the baseline (first) measurement. To explore if this relationship is treatment-dependent, we conducted the analysis separately for each treatment group. Given that recanalization status following thrombectomy significantly influences neurological outcomes, we verified if the relationship between inflammation and neurological outcomes depends on recanalization status. To this end, a linear mixed model was used, including IL-6 levels, treatment, the interaction between treatment and IL-6 levels, recanalization status, and the interaction between recanalization status and IL-6 levels as predictors. We also explored the relation between IL-6 and NIHSS overall, as well as change in IL-6 in relation to mRS90 overall.

### Statistical analysis of the Stroke Complications and Poor Outcomes (SCPO) in relation to treatment

To test the hypothesis that taVNS reduces the incidence of complications and poor outcomes, the total number of select complications and poor outcomes was modeled as a function of stroke severity, quantified by the average NIHSS score over the first three days. SCPO were defined as: hemorrhagic transformation, hemi-craniectomy, bradycardia, hypotension, infection, readmission, and hospital poor disposition (i.e. nursing home or long-term acute care hospital). In-hospital deaths were excluded from these analysis as all three patients who passed inpatient (in the treatment group) were due to comfort measures. Given the ordinal nature of total complications and poor outcomes, a generalized linear regression model with a Poisson link function was employed. The model includes total complications and poor outcomes as dependent variables and treatment and admission disability as predictors.

### Data availability

Data requests will be considered within reasonable request after agreement with the Steering Committee, provided data transfer complies with general data protection regulation and is approved by the respective ethical review board.

## Results

### Characteristics of participants

We randomized 40 participants; after applying all study eligibility criteria, we excluded five participants post-hoc and included 35 participants (17 treated, 18 sham) (**Figure 1).** Our cohort was biracial: Caucasian (77.1%) and Black (22.9%); had a mean age of 67.7 years (SD 13.7) and was 48.6% female. The baseline NIHSS was 16.4 (SD 6.6) for the treatment group and 15.7 (SD 4.4) for the sham group. The treated group had a higher burden of left-sided stroke (64.7% vs 44.4%), ICA occlusions (47.1% vs 27.8%), and greater time from LKN to recanalization (mean 13.2 hrs vs 9.8 hrs) when compared to the sham group, but these differences were not statistically significant. Thrombolytics were administered to just under half the cohort and thrombectomy was attempted in the majority (94%), with 82% of the treated group and 89% of the sham group achieving recanalization after thrombectomy (TICI ≥2B). The treated group had greater early edema at baseline, prior to the effects of treatment, as compared to the sham group based on cerebrospinal fluid (CSF) changes, Δ CSF −26% vs −13%, p=0.05. The mean time to first stimulation from LKN was 22.8 hrs (SD 6.5) in the treated vs 24.8 hrs (SD 6.2) in the sham group (**Table 1)**.

**Table 1.**
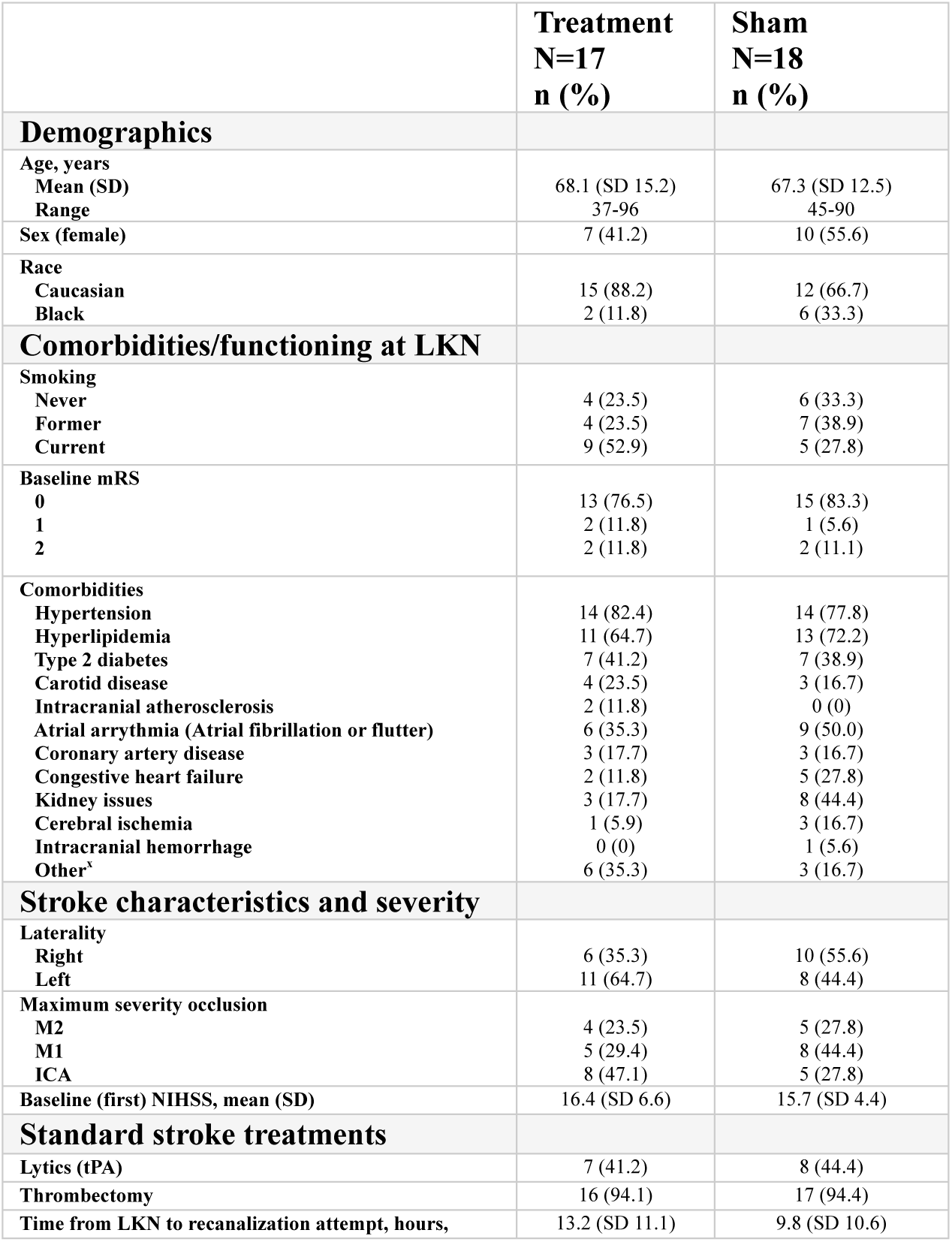
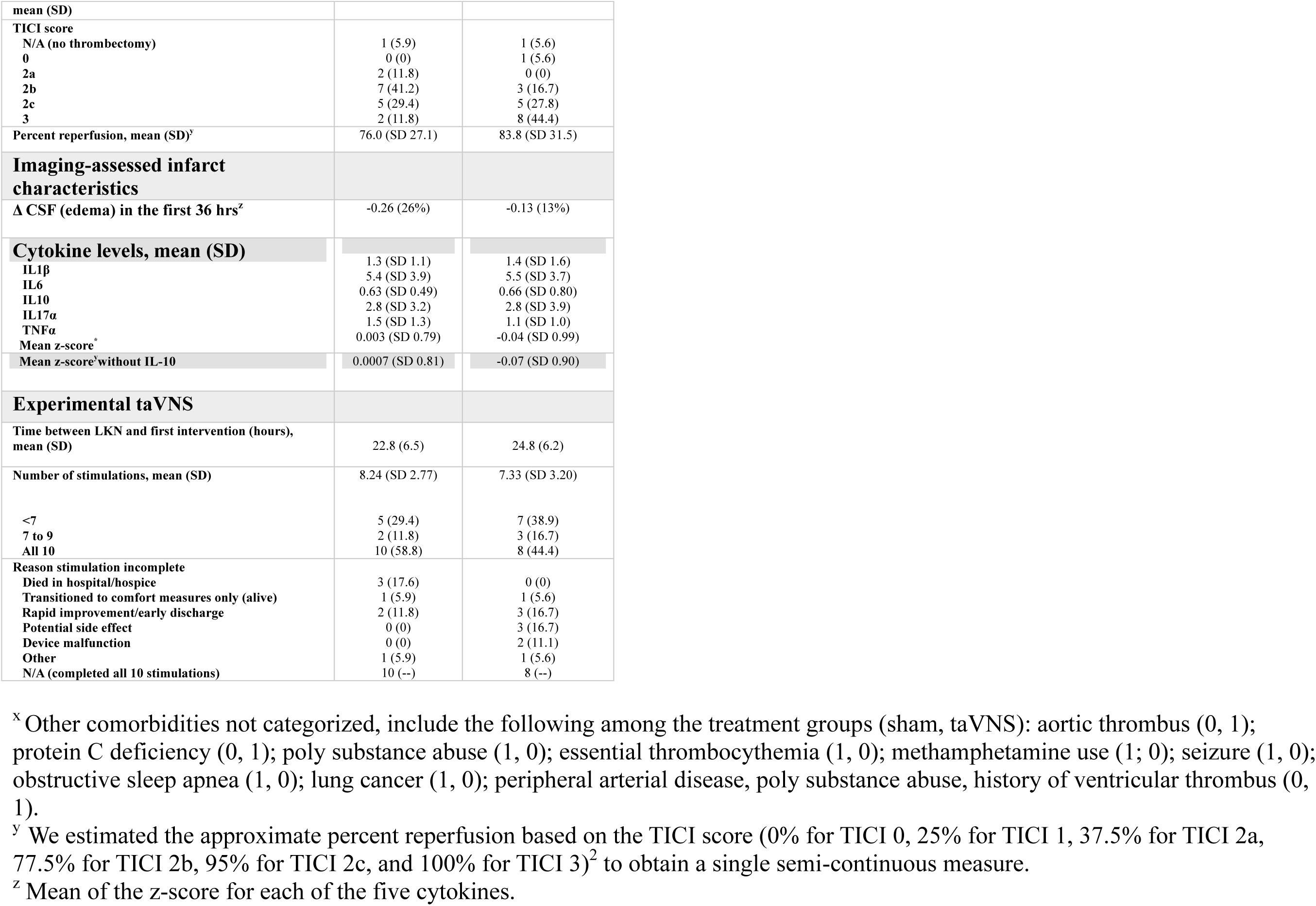
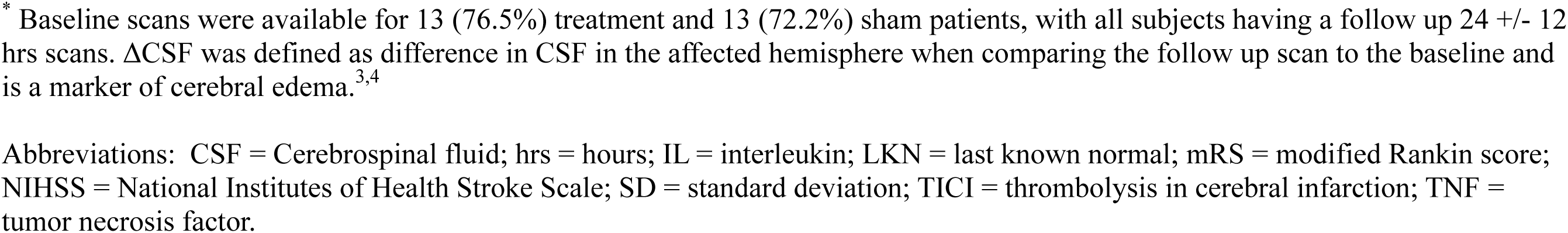
Baseline characteristics of participants and adherence to randomized treatment, by treatment group, NUVISTA.

|  | **Treatment**<br>**N=17**<br>**n (%)** | **Sham**<br>**N=18**<br>**n (%)** |
| --- | --- | --- |
| **Demographics** |  |  |
| Age, years |  |  |
| Mean (SD) | 68.1 (SD 15.2) | 67.3 (SD 12.5) |
| Range | 37-96 | 45-90 |
| Sex (female) | 7 (41.2) | 10 (55.6) |
| Race |  |  |
| Caucasian | 15 (88.2) | 12 (66.7) |
| Black | 2 (11.8) | 6 (33.3) |
| **Comorbidities/functioning at LKN** |  |  |
| Smoking |  |  |
| Never | 4 (23.5) | 6 (33.3) |
| Former | 4 (23.5) | 7 (38.9) |
| Current | 9 (52.9) | 5 (27.8) |
| Baseline mRS |  |  |
| 0 | 13 (76.5) | 15 (83.3) |
| 1 | 2 (11.8) | 1 (5.6) |
| 2 | 2 (11.8) | 2 (11.1) |
| Comorbidities |  |  |
| Hypertension | 14 (82.4) | 14 (77.8) |
| Hyperlipidemia | 11 (64.7) | 13 (72.2) |
| Type 2 diabetes | 7 (41.2) | 7 (38.9) |
| Carotid disease | 4 (23.5) | 3 (16.7) |
| Intracranial atherosclerosis | 2 (11.8) | 0 (0) |
| Atrial arrhythmia (Atrial fibrillation or flutter) | 6 (35.3) | 9 (50.0) |
| Coronary artery disease | 3 (17.7) | 3 (16.7) |
| Congestive heart failure | 2 (11.8) | 5 (27.8) |
| Kidney issues | 3 (17.7) | 8 (44.4) |
| Cerebral ischemia | 1 (5.9) | 3 (16.7) |
| Intracranial hemorrhage | 0 (0) | 1 (5.6) |
| Other<sup>a</sup> | 6 (35.3) | 3 (16.7) |
| **Stroke characteristics and severity** |  |  |
| Laterality |  |  |
| Right | 6 (35.3) | 10 (55.6) |
| Left | 11 (64.7) | 8 (44.4) |
| Maximum severity occlusion |  |  |
| M2 | 4 (23.5) | 5 (27.8) |
| M1 | 5 (29.4) | 8 (44.4) |
| ICA | 8 (47.1) | 5 (27.8) |
| Baseline (first) NIHSS, mean (SD) | 16.4 (SD 6.6) | 15.7 (SD 4.4) |
| **Standard stroke treatments** |  |  |
| Lytics (tPA) | 7 (41.2) | 8 (44.4) |
| Thrombectomy | 16 (94.1) | 17 (94.4) |
| Time from LKN to recanalization attempt, hours, | 13.2 (SD 11.1) | 9.8 (SD 10.6) |
| mean (SD) |  |  |
| **TICI score** |  |  |
| N/A (no thrombectomy) | 1 (5.9) | 1 (5.6) |
| 0 | 0 (0) | 1 (5.6) |
| 2a | 2 (11.8) | 0 (0) |
| 2b | 7 (41.2) | 3 (16.7) |
| 2c | 5 (29.4) | 5 (27.8) |
| 3 | 2 (11.8) | 8 (44.4) |
| Percent reperfusion, mean (SD)<sup>y</sup> | 76.0 (SD 27.1) | 83.8 (SD 31.5) |
| **Imaging-assessed infarct characteristics** |  |  |
| Δ CSF (edema) in the first 36 hrs<sup>z</sup> | -0.26 (26%) | -0.13 (13%) |
| **Cytokine levels, mean (SD)** |  |  |
| IL1β | 1.3 (SD 1.1) | 1.4 (SD 1.6) |
| IL6 | 5.4 (SD 3.9) | 5.5 (SD 3.7) |
| IL10 | 0.63 (SD 0.49) | 0.66 (SD 0.80) |
| IL17α | 2.8 (SD 3.2) | 2.8 (SD 3.9) |
| TNFα | 1.5 (SD 1.3) | 1.1 (SD 1.0) |
| Mean z-score<sup>*</sup> | 0.003 (SD 0.79) | -0.04 (SD 0.99) |
| Mean z-score<sup>z</sup> without IL-10 | 0.0007 (SD 0.81) | -0.07 (SD 0.90) |
| **Experimental taVNS** |  |  |
| Time between LKN and first intervention (hours), mean (SD) | 22.8 (6.5) | 24.8 (6.2) |
| Number of stimulations, mean (SD) | 8.24 (SD 2.77) | 7.33 (SD 3.20) |
| <7 | 5 (29.4) | 7 (38.9) |
| 7 to 9 | 2 (11.8) | 3 (16.7) |
| All 10 | 10 (58.8) | 8 (44.4) |
| Reason stimulation incomplete |  |  |
| Died in hospital/hospice | 3 (17.6) | 0 (0) |
| Transitioned to comfort measures only (alive) | 1 (5.9) | 1 (5.6) |
| Rapid improvement/early discharge | 2 (11.8) | 3 (16.7) |
| Potential side effect | 0 (0) | 3 (16.7) |
| Device malfunction | 0 (0) | 2 (11.1) |
| Other | 1 (5.9) | 1 (5.6) |
| N/A (completed all 10 stimulations) | 10 (--) | 8 (--) |

<sup>x</sup> Other comorbidities not categorized, include the following among the treatment groups (sham, taVNS): aortic thrombus (0, 1); protein C deficiency (0, 1); poly substance abuse (1, 0); essential thrombocythemia (1, 0); methamphetamine use (1; 0); seizure (1, 0); obstructive sleep apnea (1, 0); lung cancer (1, 0); peripheral arterial disease, poly substance abuse, history of ventricular thrombus (0, 1).
<sup>y</sup> We estimated the approximate percent reperfusion based on the TICI score (0% for TICI 0, 25% for TICI 1, 37.5% for TICI 2a, 77.5% for TICI 2b, 95% for TICI 2c, and 100% for TICI 3)<sup>2</sup> to obtain a single semi-continuous measure.
<sup>z</sup> Mean of the z-score for each of the five cytokines.
\* Baseline scans were available for 13 (76.5%) treatment and 13 (72.2%) sham patients, with all subjects having a follow up 24 +/- 12 hrs scans. $\Delta$ CSF was defined as difference in CSF in the affected hemisphere when comparing the follow up scan to the baseline and is a marker of cerebral edema.<sup>3,4</sup>
Abbreviations: CSF = Cerebrospinal fluid; hrs = hours; IL = interleukin; LKN = last known normal; mRS = modified Rankin score; NIHSS = National Institutes of Health Stroke Scale; SD = standard deviation; TICI = thrombolysis in cerebral infarction; TNF = tumor necrosis factor.

### Longitudinal change in cytokines in relation to treatment group

Across individual cytokines and the z score-based summary measures, differences in the pattern of change over time between the treatment and sham groups were suggested in both our primary analysis and the sensitivity analysis utilizing a time-varying exposure (treatment) variable (**Table 2**, **Figure 2, and Supplemental Figure 1)**. The greatest difference between the groups was observed for IL-6. Without accounting for the lack of exposure prior to randomization, the treatment and sham groups appeared much more similar than in the time-varying model. In this latter model, the two groups diverged, with the groups having U-shaped associations similar in magnitude but opposite in direction (upward U-shape for the treatment group and inverted U-shape for the sham group, p_interacton_ = 0.0001). All other cytokines studied (i.e. IL-1β, IL-10, IL-17α, and TNFα) and WBC, despite having different trajectories, did not reach statistical significance. The pattern of change in normalized pro-inflammatory cytokines over time was also different according to both assigned treatment group (p_interacton_=0.01) or time-varying treatment (p_interacton_=0.04) (**Table 2 and Figure 2)**. Looking at IL-6 specifically, levels were significantly lower in the taVNS treatment group on Day 3 compared with the sham group [t = 2.083, p = 0.045, N(taVNS)=16, N(sham) = 17, t-test, Cohen’s d = −0.726] (**Figure 2B)**. Furthermore, normalized pro-inflammatory cytokines were significantly lower in the taVNS treatment groups on Day 3, compared to the sham treatment group [t = −2.632, p = 0.013, N(taVNS)=16, N(sham) = 17, t-test, Cohen’s d = −0.917] (**Figure 2F)**, but were not significantly different on Days 1, 5, and 7.

**Figure 2.**
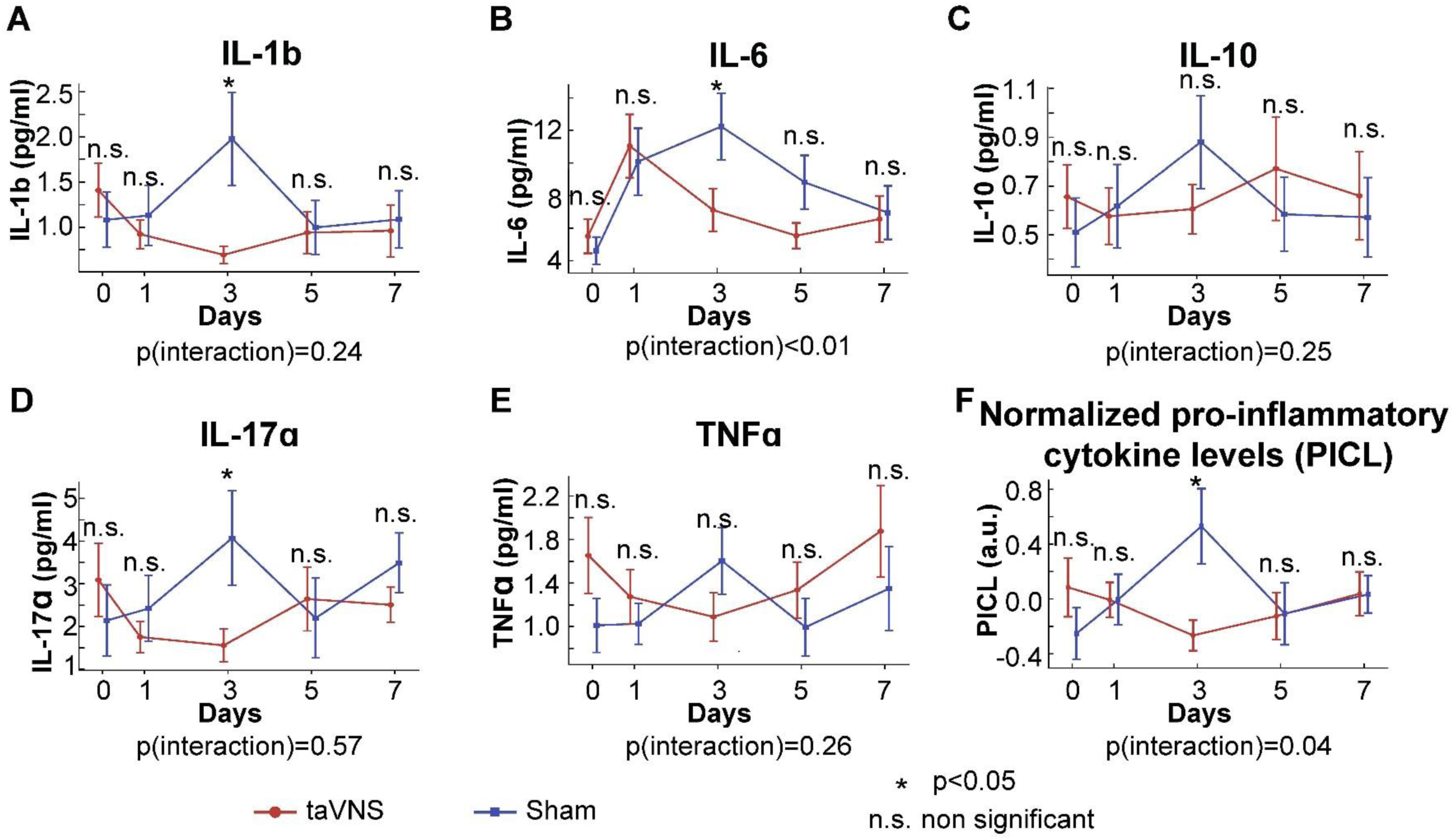
Trajectory of the different cytokine levels individually and as a composite normalized value by treatment vs sham group. P(interaction) is from the time-varying model and tests whether there is a statistically significant difference in the cytokine level trajectory longitudinally according to randomized treatment (sham before randomization and assigned treatment group for all points after randomization). The P(interaction) in the time varying model showed a significant difference amongst the trajectory IL-6 cytokine levels over days 0-7 when comparing treatment vs sham. **A,B, D**. IL-1b, IL-6, and IL-17a levels for taVNS and sham treatment groups were significantly lower in the taVNS treatment group on day 3 compared with the sham treatment group, e.g. IL-6 [t = 2.083, p = 0.045, N(taVNS)=16, N(sham) = 17, t-test]. The error bars in **A-F** represent standard errors. **F**. Normalized pro-inflammatory cytokine (TNF-a, IL-1β, IL-17a, and IL-6) score over days 0-7 for taVNS and sham treatment groups (randomized on approximately day 1 and treated through day 5, death, or discharge, whichever came earliest). Normalized pro-inflammatory cytokine levels were significantly lower in the taVNS treatment group on day 3 compared with the sham treatment group [t = 2.632, p = 0.013, N(taVNS)=16, N(sham) = 17, t-test]. The P(interaction) in the time varying model showed a significant difference amongst the trajectory of the normalized pro-inflammatory cytokine levels when comparing treatment vs sham.

**Table 2.**
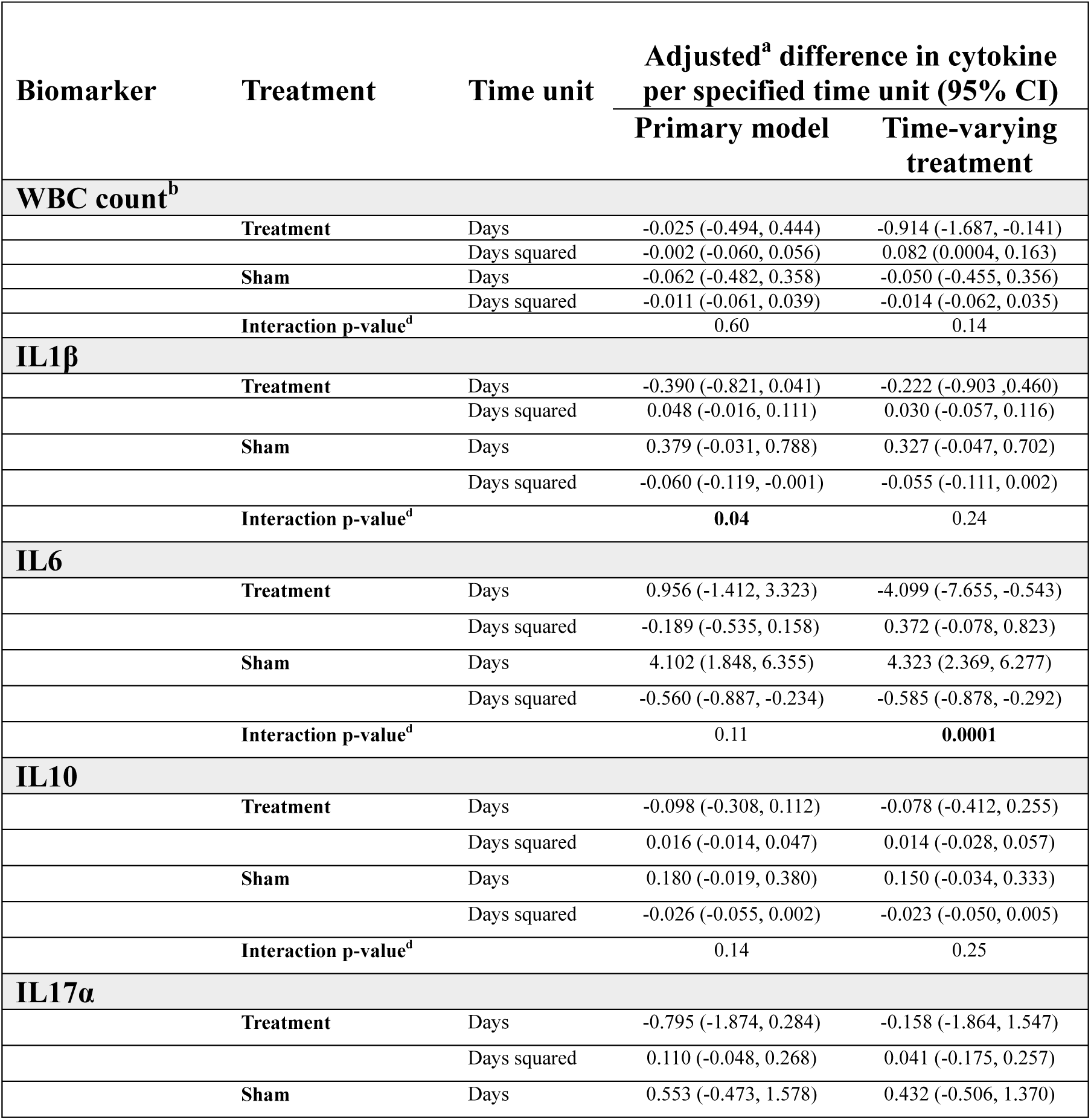
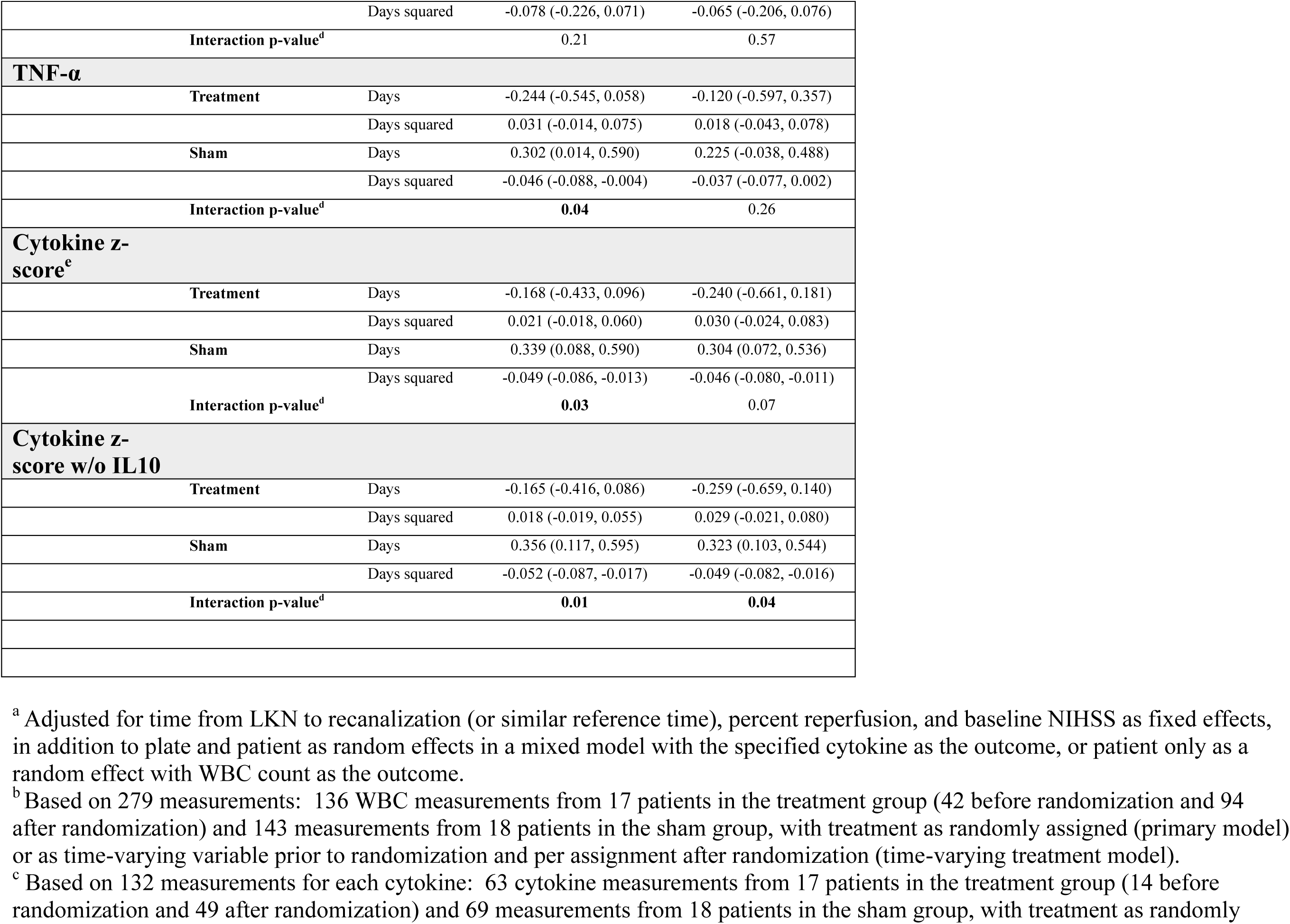
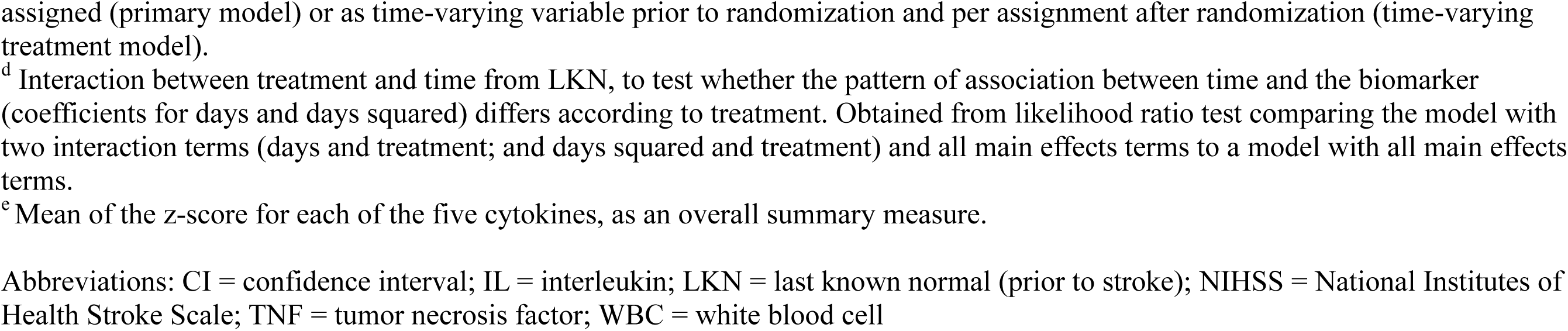
Blood biomarkers of inflammation in relation to time from last known normal, by treatment.

### IL-6 and clinical outcomes

Across combined treatment groups, changes in IL-6 demonstrated a correlation with Day 90 mRS scores (95% CI for β_(IL-6) = [0.061, 0.177], p <0.001, z-test). For the acute inpatient recovery period, a statistically significant relationship was also identified between IL-6 and NIHSS scores (F-statistic = 5.276, p = 0.023, *R*^2^ = 0.040), with an IL-6 coefficient of 0.252 (p = 0.023, t-test, n = 130).

### Change in IL-6 and change in NIHSS by treatment group

Changes in NIHSS scores were positively associated with changes in IL-6 levels, indicating reduced pro-inflammatory cytokine levels were associated with improving neurological outcomes (slope = 0.358, 95% CI = [0.055, 0.661], t = 2.370, p = 0.021, n = 57, t-test). The regression performed separately for the two treatment groups showed significant positive correlation between changes in IL-6 levels and changes in NIHSS scores in the taVNS group (slope = 0.798, 95% CI = [0.077, 1.518], t = 2.286, p = 0.031, n = 26, t-test) and borderline significant relationship in the sham group (slope = 0.330, 95% CI = [−0.029, 0.690], t = 1.881, p = 0.070, n = 31, t-test). That is, for the taVNS treatment group, a 1 pg/mL reduction in IL-6 over 3 to 5 days was associated with a reduction of 0.80 points in NIHSS (**Figure 3A)**. The model incorporating subject random effect largely confirmed this finding; the association was significant in the taVNS treatment group and non-significant in the sham treatment group (**Supplemental Figure 2).**

**Figure 3.**
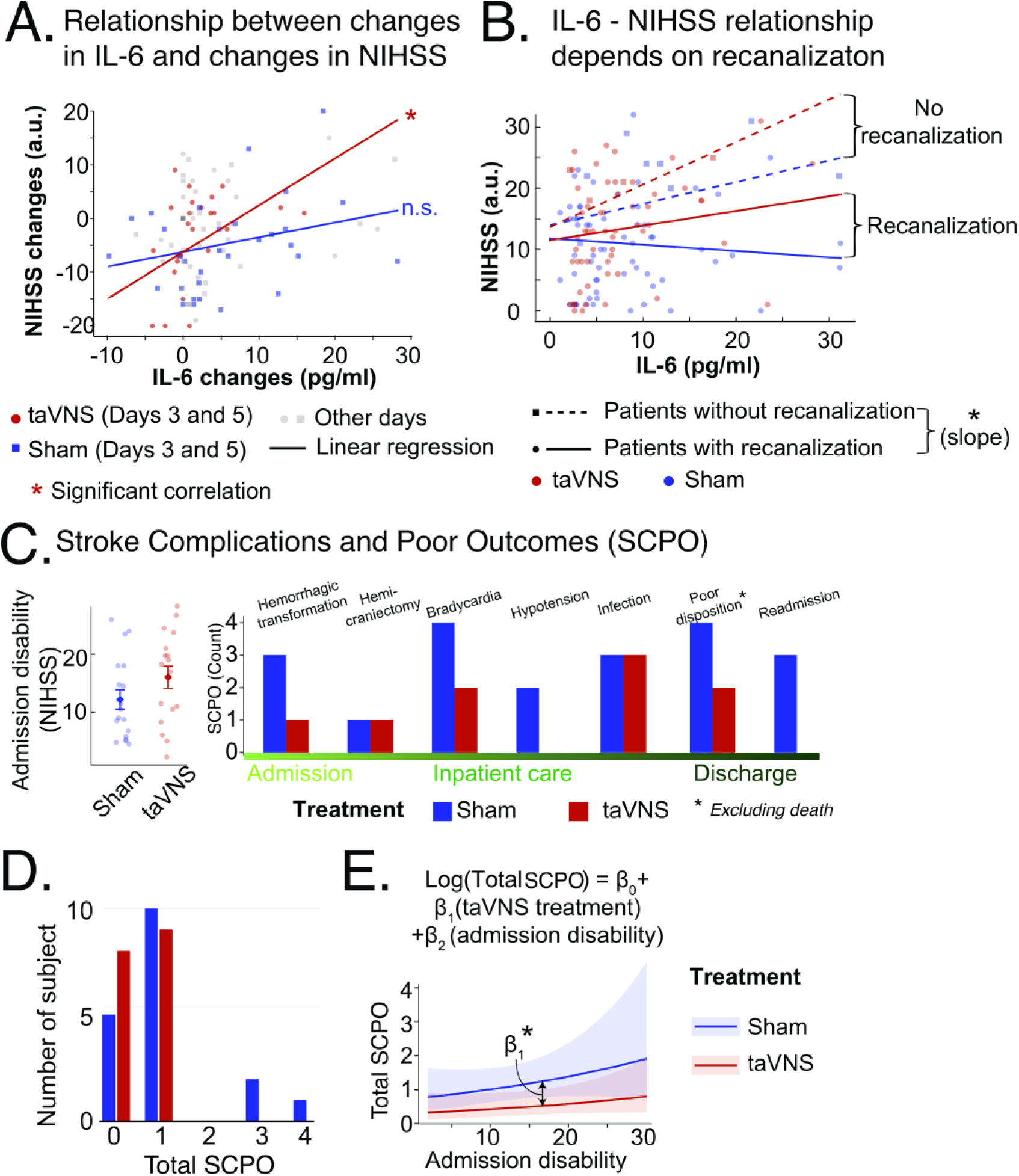
**A**. Changes in NIHSS scores were significantly positively associated with changes in IL-6 levels in the taVNS treatment group, suggesting that reduced pro-inflammatory cytokine levels were associated with improving neurological outcomes (slope = 0.798, 95% CI = [0.077, 1.518], t = 2.286, p = 0.031, n = 26, t-test). The relationship between changes in NIHSS scores and changes in IL-6 levels was not significant in the sham group (slope = 0.330, 95% CI = [−0.029, 0.690], t = 1.881, p = 0.070, n = 31, t-test). **B**. The positive association between change in IL-6 and change in NIHSS was greater among patients without recanalization. A 1 pg/mL increase in IL-6 was associated with an additional 0.46-point increase in NIHSS scores in patients who did not achieve recanalization compared with patients who did (z=-2.015, p=0.044, 95%CI = [0.012, 0.898]). **C-E.** Stroke complications and poor outcomes (SCPO) in the two treatment groups. **C.** The mean initial stroke severity was higher in the taVNS group measured by average NIHSS for Days 0 – 3. The right figure shows the occurrence of stroke complications and poor outcomes for the two treatment groups, while excluding patients who died inpatient due to comfort measures as they did not have time to develop inflammation related complications. **D.** Distribution of total stroke complications and poor outcomes by treatment group. **E**. The relationship between total SCPO and initial stroke severity for the two treatment groups. The 95% CI for the slope 𝛽_2_ is [−0.018, 0.081] (z = 1.264, p = 0.206). Total SCPO was significantly higher in the sham treatment group (95% CI for the difference in intercept between the two treatment groups = [−1.680, −0,058], z = −2.097, p = 0.036).

### The relationship between IL-6 and NIHSS by recanalization status

When we analyzed how recanalization status modulates the relationship between pro-inflammatory cytokine IL-6 and NIHSS scores by including recanalization status and its interaction with IL-6, our analysis revealed that the positive association between change in IL-6 and change in NIHSS was greater among patients without recanalization. A 1 pg/mL increase in IL-6 was associated with an additional 0.46-point increase in NIHSS scores in patients who did not achieve recanalization compared with patients who did (z=-2.015, p=0.044, 95%CI = [0.012, 0.898], z-test (**Figure 3B**). In this model, i.e., while accounting for a possible interaction between IL-6 and recanalization status, patients in the taVNS treatment group showed a trend toward a stronger IL-6/NIHSS relationship, although this difference did not reach statistical significance (95%CI of the slope difference = [0.002, 0.685], p = 0.051, z-test, t = 1.951).

### Change in NIHSS by laterality

During hospitalization, NIHSS improvement rates were initially similar between treatment groups, even when restricting to the first five days in sensitivity analysis, as shown in **Supplemental Table 2**. A significant three-way interaction between treatment group, time, and laterality was observed, with more rapid improvement noted in treated vs. sham patients with left sided strokes, i.e., ipsilateral to the site of stimulation, but not right sided strokes. This three-way interaction remained significant when we explored the effect of adding a parallel three-way interaction for occlusion type. The laterality effect noted above was also seen in cytokines generally and IL-6 most clearly demonstrated a difference between treatment groups for left-side strokes (p_interaction_ = 0.0005) (**Supplemental Table 3)**.

### Stroke Complications and Poor Outcomes in relation to treatment

No adverse events directly attributable to taVNS were noted. Primary cardiac events were observed more frequently in the sham group, including bradycardia (22.2% vs 11.8%) and hypotension (11.1% vs 0%). Hemorrhagic transformation was documented more frequently in the sham group (16.7% vs 5.9%). Other conditions that could be associated with inflammation occurred at similar rates between groups (sham 27.8% vs treatment 23.5%), including infections (three in each group) and hemicraniectomy (one patient in each group), but none of these were statistically significant. While in-hospital mortality was higher in the treatment group (17.6% vs 0%), at the mRS 90 day mark, the treatment group had three deaths (17.6%), and the sham group had two deaths (11.1%), and none of these findings were statistically significant. Those patients who passed in the treatment group were statistically significantly older than those patients who did not die (passed age 84.7 (SD 7.7) vs alive age 64.5 (SD 3.6), p = 0.03), were on the trial on average only 2.17 days, and all passed due to the family/patient’s wishes being congruent with comfort measures only. In addition, overall poor dispositions were similar in the two groups and good discharge dispositions were achieved in comparable proportions (treatment 70.6% vs sham 77.8%). Hospital readmissions were recorded only in the sham group (16.7%). Although none of these individual differences reached statistical significance (all p≥0.60), we noted a trend when taken in aggregate. Despite having higher stroke severity and larger infarct volumes, patients treated with taVNS appeared to have less morbidity when excluding patients who died early after admission (**Figure 3 C-E).** The generalized linear regression modeling total stroke complications and poor outcomes as a function of treatment and admission disability showed that surviving patients in the taVNS treatment group experienced significantly fewer complications and poor outcomes than the surviving sham group (incidence rate ratio = 0.42, 95% CI: [0.19, 0.94], z = −2.097, p = 0.036) (**Figure 3 and Table 3).** When deaths were included in the analysis, surviving patients in the taVNS treatment group experienced 51% fewer complications and poor outcomes than the surviving sham group, but the difference was non-significant ( 95% CI of the incidence rate ratio = [0.24, 1.01], z = −1.921, p = 0.055).

**Table 3.** Adverse Events and Secondary Outcomes.

|  | **Treatment**<br>**N=17**<br>**n (%)** | **Sham**<br>**N=18**<br>**n (%)** | **Unadjusted**<br>**p-value** |
| --- | --- | --- | --- |
| **Cardiac Events** |  |  |  |
| Bradycardia | 2 (11.8) | 4 (22.2) | $p_{\text{exact}} = 0.66$ |
| Hypotension | 0 (0.0) | 2 (11.1) | $p_{\text{exact}} = 0.49$ |
| **Adverse Events** |  |  |  |
| Infection | 3 (17.7) | 3 (16.7) | $p_{\text{exact}} = 1.00$ |
| Hemicraniectomy | 1 (5.9) | 1 (5.6) | $p_{\text{exact}} = 1.00$ |
| Hemorrhagic transformation | 1 (5.9) | 3 (16.7) | $p_{\text{exact}} = 0.60$ |
| **Other secondary outcomes** |  |  |  |
| Early discharge due to improvement | 2 (11.8) | 3 (16.7) | $p_{\text{exact}} = 1.00$ |
| Discharge disposition <sup>†</sup> | | | $p_{\text{trend}} = 0.24$ |
| Good | 12 (70.6) | 14 (77.8) |  |
| Poor | 2 (11.8) | 4 (22.2) |  |
| Died before discharge** | 3 (17.7) | 0 (0) |  |
| Readmitted to hospital | 0 (0) | 3 (16.7) | $p_{\text{exact}} = 0.23$ |
| mRS at 30 days,*** mean (SD) | 3.63 (1.82) | 2.83 (1.92) | $p_{\text{trend}} = 0.21$ |
| 0 | 1 (6.3) | 2 (11.1) |  |
| 1 | 1 (6.3) | 5 (27.8) |  |
| 2 | 3 (18.8) | 0 (0) |  |
| 3 | 1 (6.3) | 3 (16.7) |  |
| 4 | 5 (31.3) | 4 (22.2) |  |
| 5 | 2 (12.5) | 3 (16.7) |  |
| 6 | 3 (18.8) | 1 (5.6) |  |
| Missing | 1 (–) | 0 (–) |  |
| mRS at 90 days,*** mean (SD) | 3.27 (2.15) | 2.78 (2.07) | $p_{\text{trend}} = 0.50$ |
| 0 | 2 (13.3) | 4 (22.2) |  |
| 1 | 2 (13.3) | 2 (11.1) |  |
| 2 | 2 (13.3) | 1 (5.6) |  |
| 3 | 1 (6.7) | 4 (22.2) |  |
| 4 | 3 (20.0) | 3 (16.7) |  |
| 5 | 2 (13.3) | 2 (11.1) |  |
| 6 | 3 (20.0) | 2 (11.1) |  |
| Missing | 2 (--) | 0 (--) |  |
| Length of stay among patients discharged alive<br>and not to hospice, days, mean (SD) | 8.01 (4.16) | 8.91 (5.34) | $p_{\text{trend}} = 0.59$ |

† Good disposition includes home or inpatient rehabilitation facility; poor disposition includes long term care hospital, skilled nursing facility.
\*\* We excluded inpatient death from poor disposition to restrict to those patients who would have had time for an inflammatory response to be the cause of the death. Those patients who passed in the treatment group were statistically significantly older than those patients who did not die (passed age 84.7 (SD 7.7) vs alive 64.5 (SD 3.6), $p = 0.03$ ), were on the trial on average only 2.17 days, and all passed due to the family/patient's wishes being congruent with comfort measures only.
\*\*\* Percentages shown among those without missing data. mRS at 30 days excludes one participant from the treatment group lost to follow-up; mRS at 90 days also excludes one additional participant from the treatment group. The two patients who died in the sham group, one bounced back due to atrial fibrillation with rapid ventricular response leading to cardiac arrest and the other patient's cause of death was not able to be identified due to lack of access to their external medical record.

## Discussion

The NUVISTA trial demonstrated that taVNS, when added to standard medical care, safely and effectively modulates the post-AIS inflammatory response in patients with anterior circulation LVO strokes. Patients receiving taVNS within 36 hours of symptom onset showed significantly faster reductions in inflammatory cytokines, most notably in IL-6, in the first days following the stroke onset after initiation of treatment. In addition, a reduction in IL-6 significantly correlated with a reduction in neurologic deficit and may be more pronounced in patients with persistent LVO or unsuccessful recanalization via thrombectomy. This randomized pilot trial demonstrated that taVNS may aid in mitigating the inflammatory sequelae associated with AIS and may reduce its associated neurologic complications and morbidity.

Although the study was not powered to detect differences in functional neurologic outcomes in this study we have confirmed that elevated pro-inflammatory cytokines, in particular IL-6, are associated with worsened functional outcomes, as previously reported in the literature.^6–10,42–44^ Furthermore, the taVNS treatment and sham groups were not balanced: the taVNS treatment group had more proximal LVO locations, larger infarct sizes, longer times from LKN to recanalization, worse reperfusion scores, and more early edema development than the sham group. Despite these baseline imbalances between groups, we still saw an effect on inflammatory biomarkers, highlighting the potential robustness of this intervention. Further, when clinical outcomes were assessed relative to the change in IL-6, there were indeed differences between the taVNS-treated and sham groups, with treated surviving patients experiencing significantly fewer complications and poor outcomes compared to the sham group.

Our findings demonstrate that taVNS treatment was associated with a greater correlation between reduced IL-6 levels and improved NIHSS scores, with every 1 pg/mL decrease in IL-6 corresponding to a reduction of 0.798 points in NIHSS scores (95% CI [0.077, 1.518]), as compared to 0.330 (95% CI [−0.029, 0.690] reduction in the sham group. In addition, the overall relationship between change in IL-6 and change in NIHSS may be stronger in patients who did not have successful recanalization, suggesting that taVNS may offer additional therapeutic benefit in cases where mechanical thrombectomy fails to restore blood flow. Taken together, these findings suggest a potential role for taVNS as an adjunctive therapy and further exploration in patients where standard recanalization strategies are unsuccessful or only partially effective may be valuable

The mechanisms of action of VNS and its impact on clinical outcomes may be pleiotropic. It is postulated that taVNS leads to immunomodulation through neural pathways associated with the cholinergic anti-inflammatory pathway, parasympathetic nervous system, and hypothalamic-pituitary-adrenal (HPA) axis. Immunogenic stimuli activate vagal afferents terminating primarily in the dorsal vagal complex. Ascending projections from the dorsal vagal complex reach the paraventricular nucleus (PVN) and rostral ventromedial medulla (RVM). The arrival of these incoming signals generates action potentials that travel from the brainstem to the spleen and other organs. This culminates in T cell release of acetylcholine, which interacts with α7 nicotinic acetylcholine receptors (α7 nAChR) on immunocompetent cells to inhibit cytokine release in macrophages.^45^ This is likely the mechanism explaining this study’s reduction in inflammatory cytokines. Transcutaneous auricular VNS can also positively impact cardiovascular function through the sympathetic nervous system or efferent vagus nerve.^46^ Furthermore, it can reduce blood-brain barrier permeability, cortical spreading depressions, and improve regionalized plasticity.^46,47^ In aggregate, these pleiotropic effects may play a role in reducing stroke complications and improving outcomes.

Interestingly, we saw differential effects of the stimulation depending on the laterality of the stroke. In this study, we exclusively stimulated the left vagus nerve, which is a widely accepted side, to avoid stimulation of cardiac centers.^48^ Despite this practice, prior reports suggest that stimulation may induce a more robust brain activation in the ipsilateral hemisphere.^39,40^ Although the differences we saw may be due to a limited-size study group, this should be explored further in future trials.

This trial has notable strengths, including its randomized, blinded design and comprehensive biomarker analysis. However, several important limitations warrant discussion. First, the small sample size introduces the possibility that our findings, particularly the clinical outcomes, may be subject to chance variation, though the consistency of cytokine changes provides some reassurance. Second, as a single-center study not powered to detect clinical differences, these results require validation through larger, multicenter trials. Third, fundamental questions remain about optimal therapeutic parameters, including stimulation settings, treatment duration, and most relevant physiologic and biomarker endpoints when utilizing taVNS. This is highlighted in our trial by the Day 5 to 7 rises in some cytokine levels, which may be a rebound effect of stopping treatment, although an additional explanation could be that our sample size decremented at those time points and stroke patients who were still in the hospital at that time tended to be sicker. Finally, the focus on large vessel occlusion strokes limits the generalizability of these findings to other stroke subtypes. These limitations highlight critical areas for future investigation while providing a foundation for larger-scale studies of taVNS in acute ischemic stroke.

In conclusion, the NUVISTA trial demonstrates that taVNS safely modulates post-AIS inflammation in anterior circulation large vessel occlusion, with reductions in IL-6 correlating with improved NIHSS in the days following stroke. These findings support further investigation of taVNS as a potential adjunctive therapy through larger, multicenter trials.

## Supporting information

NUVISTA Supplements

## Acknowledgements

This work was supported, in part, by the Bursky Center for Human Immunology and Immunotherapy Programs at Washington University, Immunomonitoring Laboratory. We would like to acknowledge Dr. Diane Bender, the lead scientist/lab manager of the Immunomonitoring laboratory, for her help with cytokine assessment. We would like to acknowledge the NUVISTA coordinators: Angela Birke and Dora Murphy; and Karen McDonell for her aid with data cleaning and storage.

## Sources of Funding

This trial was supported by: a Clinical and Translational Research Funding Program (CTRFP) award UL1TR002345 (for OJLS); NIH/NINDS R37 NS110699 (for JML); NIH/NINDS R01NS121218 (for RD); and NIH/NINDS K23NS136843 (for JG).

## Disclosures

Dr. Leuthardt reports stock ownership in Neurolutions, Face to Face Biometrics, Caeli Vascular, Acera, Sora Neuroscience, Inner Cosmos, Inflexion Vascular, Aurenar, Petal Surgical, Cordance Medical, and Silent Surgical. They serve as a consultant for Monteris Medical, E15, and Neurolutions. They have received licensing fees from intellectual property for Neurolutions, Caeli Vascular, and Inner Cosmos. Additionally, they have licensing/product development agreements or receive royalties for inventions/IP from Intellectual Ventures, Sora Neuroscience, Inner Cosmos, Neurolutions, and Aurenar. Washington University owns equity in Neurolutions. Dr. Anna Huguenard reports stock ownership in Aurenar and may receive royalties for interventions/IP from Aurenar. Kara Donovan reports stock ownership in Aurenar and may receive royalties for interventions/IP from Aurenar. Dr. Opeolu Adeoye reports stock ownership in Sense Diagnostics, Inc (Founder) and may receive royalties for interventions/IP. Dr. Jin-Moo Lee, Dr. Osvaldo J. Laurido-Soto, Dr. Susane Searles Nielsen, Dr. James Giles, Dr. Rajat Dhar, and Gansheng Tan do not have related financial disclosures. At no time was this trial funded, designed, nor supported by industry.

## Nonstandard abbreviations and acronyms

ANOVA: Analysis of Variance
AIS: Acute Ischemic Stroke
CRP: C-Reactive Protein
CT: Computerized Tomography
Hz: Hertz
ID: Identification Number
IFN: Interferon
IL: Interleukin
LKN: Last Known Normal
LVO: Large Vessel Occlusion
mA: milliampere
mRS: Modified Rankin Scale
MRI: Magnetic Resonance Imaging
NIHSS: National Institutes of Health Stroke Scale
taVNS: Transcutaneous Auricular Vagus Nerve Stimulator
TGF: Transforming growth factor
TICI: Thrombolysis in Cerebral Infarction Score
TNF: Tumor Necrosis Factor
μs: microseconds
VNS: Vagus Nerve Stimulation
WBC: White Blood Cell

## Notes

### Clinical Trial

NCT05390580

### Author Declarations

The study was approved by the Washington University Human Research Protection Office. All participants provided written informed consent prior to randomization. The study was registered and approved by ClinicalTrials.Gov ID: NCT05390580.

