## Supplementary material for "Transcutaneous Auricular Vagus Nerve Stimulation Reduces Inflammatory Biomarkers and May Improve Outcomes after Large Vessel Occlusion Strokes: Results of the Randomized Clinical Trial NUVISTA": NUVISTA Supplements

**Supplemental Methods.**

**Supplement Information 1.**

*Study protocol.*

Patients enrolled in the trial will be randomized to treatment with or without electrical stimulation via an auricular, transcutaneous vagus nerve stimulator. All patients are fitted with a portable Soterix^©^ device, a transcutaneous electrical nerve stimulation unit, and are connected to two ear electrodes, applied to the left ear during treatment periods. For taVNS treatment, these ear electrodes are placed along the concha of the ear, while in sham treatments the electrodes are placed along the ear lobe to avoid stimulation of the auricular vagus nerve from tactile pressure alone in the absence of current. Stimulation parameters were selected based on prior studies that sought to maximize vagus somatosensory evoked potentials while avoiding perception of pain. Stimulation parameters will be 20 minutes duration, frequency of 20 Hz, 250µs pulse width, and an intensity of 0.5mA. Sham treatments will involve no electrical current to the auricular branch of the vagal nerve. The site of stimulation will be inspected daily before and after treatment to ensure there is no redness or irritation at the site. Although the Soterix^©^ device is an FDA approved product, we utilized it off-label under a Non-Significant Risk (NSR) Device clearance approved by our institution’s IRB. Blood samples are collected on day 1, 3, 5, and 7, for complete blood count with differential. For measurement of inflammatory cytokines, samples are centrifuged, aliquoted, and stored in a deep freezer at -80 °C until ready for processing. Luminex^®^ multiplex assays are then run by the Bursky Center for Human Immunology & Immunotherapy Programs (CHiiPs) core lab at Washington University School of Medicine. Thawed plasma will be analyzed in duplicate with multiplex kits (Thermofisher Scientific, Waltham, MA) for the following pro-inflammatory cytokines: IL-1β, IL-6, IL-10, IL-17, and TNF-α. The concentration of each antigen is calculated by plotting the expected concentration of the standards against the multiplex fluorescent immunoassay generated by each standard. A 4-parameter logistic regression will be used for the best-fit curve. Protein concentration will be reported as pg/mL.

*Randomization and blinding.*

Dr. James Giles generated the random allocation sequence. Patients were randomized in a 1:1 ratio to either the stimulation or sham treatment group using SAS software (Version 9.4, SAS Institute, Cary, NC). A total of 80 patients were assigned to one of two groups based on a computer-generated randomization sequence using the RANUNI function with a fixed seed (43523483) to ensure reproducibility. The generated sequence was then sorted, and the first 40 patients were allocated to the stimulation group, while the remaining 40 were assigned to the sham group. Our coordinators, Dora Murphy, Angela Birke, Isabella Xu, and Dr. Osvaldo J. Laurido-Soto enrolled the participants. The member of the investigation team who generated the randomization were not involved in enrolling participants, assigning them to interventions or outcomes assessments; hence, interventions assignment was always concealed to the rest of the investigation team. Post assignment participants, care providers, and those assessing outcomes were blinded to the participants group.

*Changes post-trial initiation.*

Post trial initiation we expanded the recruitment window from 24 hrs up to 36 hrs to be able to pragmatically recruit patients based on coordinator availability. This time window was deemed acceptable as the inflammatory effects on average occur up to Day 5. We also expanded our blood sample collection to include day 7 to capture a longer trajectory of events. We intended to initially recruit 80 patients (N = 40 per group), due to recruitment issues, cost, and that based on our initial power analysis we expected to see differences in cytokines with an N = ~40 total, we stopped recruitment at N = 40 patients.

*Diverse population enrollment and diverse steering committee.*

Although no specific methodology was used to ensure a diverse population was recruited, based on our geographic location, over 50% of our participants were female and 23% were black, ensuring a diverse population. Furthermore, based on our academic setting, our steering committee and authors were from diverse backgrounds, despite no formal methodology to ensure this.

*Data management and study approvals.*

The study PI (OJLS) and Co-PIs (JML, EL) had full access to all the data in the study and takes responsibility for its integrity and supervision of the data analysis. The study was approved by the Washington University Human Research Protection Office. All participants provided written informed consent prior to randomization. The study was registered and approved by “ClinicalTrials.Gov” ID: NCT05390580.

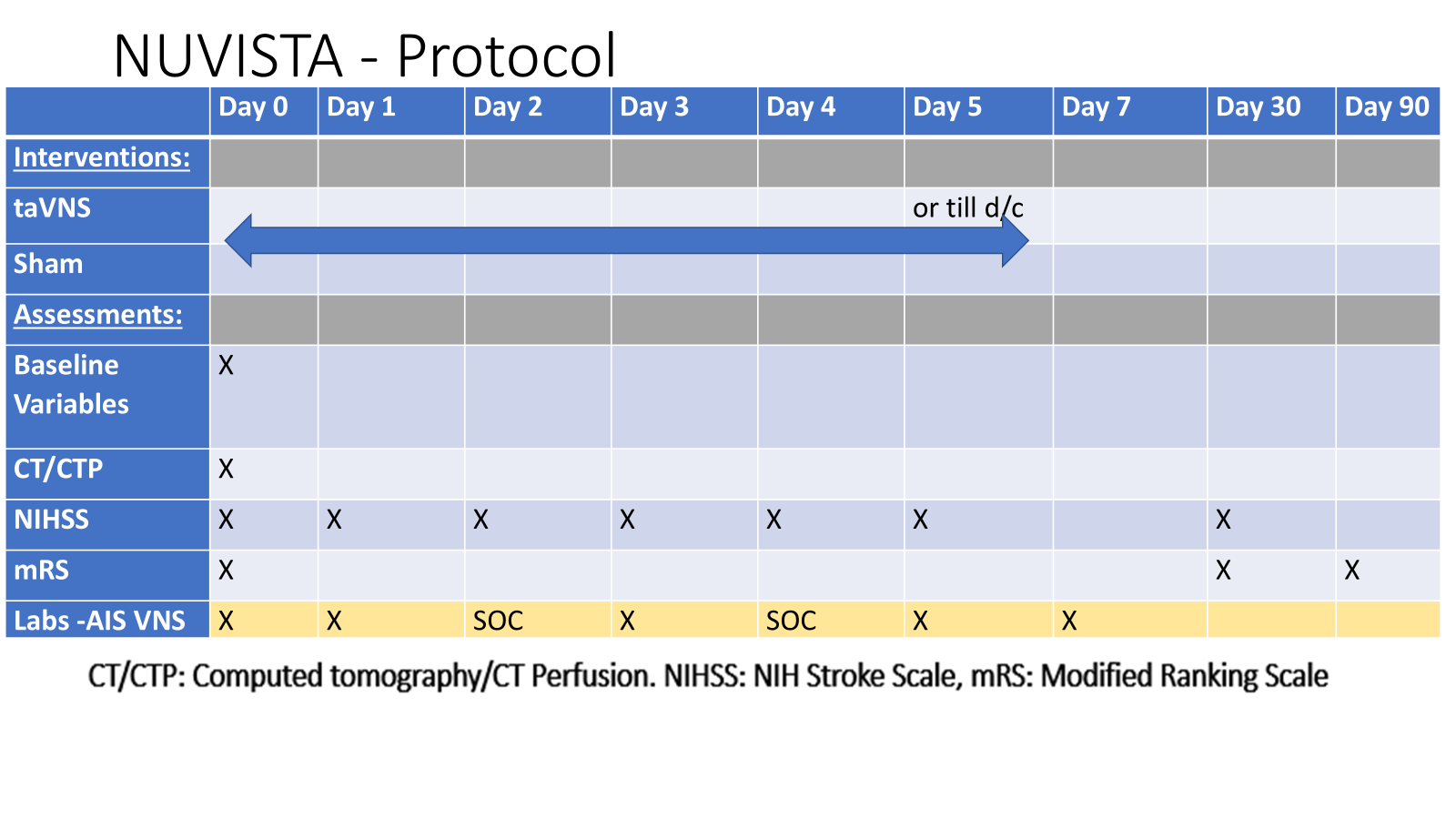

**Supplemental Information 2. Consort checklist**

| **Visit #** | **Action** | **Notes** |
| --- | --- | --- |
| **Baseline Visit** |  |  |
| 10ml | **Spin** | **Plasma – 0.5ml** |
| **24hr Visit** |  |  |
| 10ml | **Spin** | **Plasma – 0.5ml** |
| **72h Visit** |  |  |
| 10ml | **Spin** | **Plasma – 0.5ml** |
| **120hr visit** |  |  |
| 10ml | **Spin** | **Plasma- 0.5ml** |
| **168h Visit** |  |  |
| 10ml | **Spin** | **Plasma- 0.5ml** |

**Supplement Information 3. NUVISTA Blood Collection and Processing**

**Notes**

-It is important to take steps to prevent hemolysis in these samples. A vacutainer is recommended. If a needle is used, a 21 gauge needle is recommended

-After collection, gently mix the blood by inverting the tube 8 to 10 times. Store vacutainer tubes upright at 4ºC until centrifugation. Samples may be carried/stored on ice to the lab, but for no more than 2 hours. Blood samples should be centrifuged **within two hours** of blood collection.

-Gloves must be worn at all times when handling specimens. This includes during removal of the rubber stopper from the blood tubes, centrifugation, pipetting, disposal of contaminated tubes, and clean-up of any spills

**Spinning**

1) Place tubes in centrifuge, taking care to use alternate tubes to balance.
2) Program to: **10 min, 4 degrees C, 2000g (rpm) speed**, brake off.
3) Wait until it COMPLETELY stops spinning before opening.
4) After centrifugation, plasma layer will be at the top of the tube. Mononuclear cells and platelets will be in a whitish layer, called the “buffy coat”, just under the plasma and above the red blood cells.
5) Carefully collect the plasma layer with an appropriate transfer pipette. Close the caps tightly and label with the patient study number and date. **This process should be completed within 1 hour of centrifugation, which should be a total of no more than 2 hours after collection.**

**Freezing**

-Try to get in and out of the freezer as quick as possible. If open for too long, the freezer may lock to bring temperatures back down (usually about 15 min).

-Place all aliquots upright in a specimen box or rack in an -80ºC or colder freezer. All specimens should remain at -80ºC or colder prior to processing at the CHiiPs core.

**Notes**

- Please document all necessary information in the appropriate columns located on the “NUVISTA Blood Processing Documentation” This includes:

1. Date and time of blood collection

2. Number and volume of aliquots prepared

3. Date and time into -80ºC

4. Any variations or deviations from the SOP, problems, or issues

**Materials**

Simplified verbiage:

The NUVISTA blood was collected in 10cc EDTA vacutainer tubes (lavender top). They were placed on wet ice within 5min of collection and stayed on wet ice until centrifugation. We tried to process the samples within 1 hour of collecting, though a few of the samples may have been processed slightly over an hour (infrequent; if nurse needed to draw during or before intervention).

**Supplement Information 4. Methods Supplement and Trial Power Calculation**

*Assessment of blood biomarkers of inflammation.*

The cytokines were analyzed simultaneously using Luminex® multiplex assays by a core lab at Washington University School of Medicine. Thawed plasma was analyzed in duplicate with multiplex kits (Thermofisher Scientific, Waltham, MA) for the pro-inflammatory cytokines. The concentration of each antigen was calculated by plotting the expected concentration of the standards against the multiplex fluorescent immunoassay generated by each standard. A 4-parameter logistic regression was used for the best-fit curve. Protein concentrations were reported as pg/mL. For cytokines below the lower limit of quantification/limit of detection, one-half of that value was assigned for the respective cytokine.^1^

*Statistical analysis of rate of change in WBC, cytokines, and neurological outcomes in taVNS vs. sham treatment.*

Longitudinal data were analyzed using mixed effects models per the FDA,^2^ with person as a random effect, given multiple records per person (215 total in-hospital NIHSS scores, 279 WBC measures, and 132 records for each cytokine). For cytokine models, assay plate was included as an additional random effect. This approach accommodated irregularly-timed points, less restrictive missing data assumptions, and time-varying variables. To complement our primary (ITT) approach, treatment also was handled as time-varying when noted, with assignment occurring at randomization. All longitudinal models included treatment, time since last known normal (LKN), and their interaction term(s) as primary independent variables to test whether the pattern of change over time differed between the taVNS and sham groups. Time was modeled quadratically for WBC and cytokine outcomes to capture the known U-shaped associations with time^3^ when modeling these outcomes and linearly for in-hospital NIHSS scores based on panel data plots. For NIHSS a single model was constructed to estimate the rate of change (change in NIHSS per day) in each of the two treatment groups and to formally test whether these rates were different: $\Delta NIHSS \sim day+treatment+interaction \left( day, treatment \right)+covariates$using decimal days (exact assessment time). For 30-day NIHSS changes (vs. baseline) and 90-day mRS changes, we used the parallel model, with actual assessment days scaled to standardize the evaluation period to 30 or 90 days, respectively. We characterized the daily change in each WBC or cytokine outcome using a similar model expanded to include the quadratic terms: $\Delta WBC or \Delta cytokine \sim day+day squared+treatment+interaction \left( day, treatment \right)+interaction \left( day squared, treatment \right)+covariates$. Treatment-time interactions were tested using likelihood ratio tests comparing models with and without interaction term (NIHSS, mRS) or terms (WBC, cytokines), with significance at two-sided α=0.05. These analyses were adjusted for baseline covariates,^4^ including baseline NIHSS (except in NIHSS models, which included this observation), time from LKN to thrombus removal time or recanalization attempt (groin puncture time), and percent reperfusion (0-100%) based on TICI score^5^ to obtain a single semi-continuous measure. In sensitivity analyses, we also performed the analysis with a dichotomized TICI scale (reperfused ≥2B, not reperfused <2B); or restricted data to the first five days post-LKN (when most stimulations and hospitalizations had ended). Finally, we explored whether any difference in longitudinal patterns between taVNS and sham depended on laterality, occlusion type, or any of the a priori adjustment variables, i.e., whether any time-treatment interaction depended on one of these third variables.These potential interactions were assessed using three-way interaction terms in models that included all main effects and two-way interactions.

*Trial Power Calculation*.

Based on potentially recruiting 80 patients that will be randomized 1:1 to receive either taVNS (N = 40) or Sham treatment (N=40), the following calculations were made. Given a two-sided α of 0.05, this sample size will provide 90% power to detect a 0.73 standard deviation difference in each of the outcomes, and 80% power to detect this same difference for outcomes for which as many as 10 (25%) of patients have incomplete data (N=30 per group for analysis), such as due to loss to follow up due to death or attrition. Our calculations based on initial pilot data of blood and CSF inflammatory biomarkers in taVNS treated vs. not taVNS treated SAH patients,^6^ strongly suggested that we will also observe marked differences of a magnitude detectable in this study with a total sample size of N = 30-40 (N = ~20 per group), particularly for the markers assessed at 3 and 5 days.

**Supplemental Figures**

**Supplemental Figure 1**

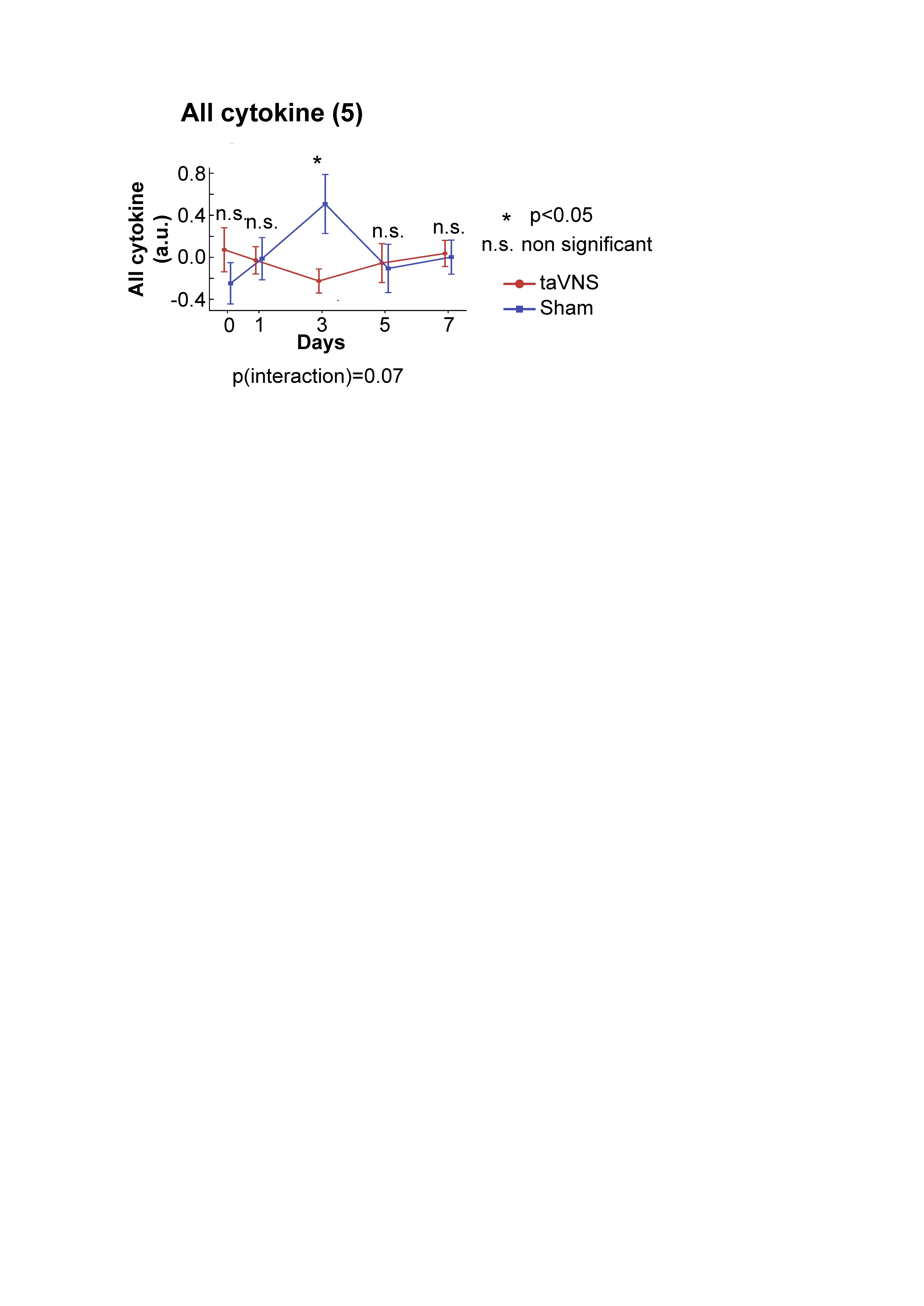

Supplementary Figure 1. Normalized cytokines (TNF-a, IL-1b, IL-10, IL-17a, and IL-6) score over days 0-7 for taVNS and sham treatment groups (randomized on approximately day 1 and treated through day 5, death, or discharge, whichever came earliest). Normalized cytokine levels were significantly lower in the taVNS treatment group on day 3 compared with the sham treatment group. The P(interaction) in the time varying model showed a borderline significant difference amongst the trajectory of the normalized pro-inflammatory cytokine levels when comparing treatment vs sham.

**Supplemental Figure 2.**

Supplementary Figure 2. The relationship between changes in IL-6 levels and changes in NIHSS was estimated using the mixed effect linear model. 95% Confidence Interval for slope in taVNS treatment group was [0.062, 0.790] (p = 0.022, t = 2.295, t-test, n = 26).

95% Confidence Interval for slope in the sham treatment group was [-0.039, 0.347] (p = 0.118, t = 1.564, t-test, n = 31).

**Supplemental Tables**

**Supplementary Table 1. NUVISTA Inclusion and Exclusions criteria**

| Inclusion | Exclusion |
| --- | --- |
| ≥ 18 years of age | Chronic Large Vessel Occlusion |
| Acute Ischemic Stroke | Pre-Morbid modified ranking scale > 2 |
| Symptom onset < 36 hours from Last Known Normal | Active Cancer, immunosuppressive or modulating therapy |
| Anterior Circulation Large Vessel Occlusion on CTA/MRA | Immune activating conditions: Chronic or severe infection, active autoimmune disease, or recent surgery/procedure |
| NIHSS ≥ 6 | Life expectancy <3 months |
|  | Heart rate < 50 or hypotension (Systolic blood pressure <80 or Mean arterial pressure < 60) |
|  | Implanted electrical device |

**Supplemental Table 2. Rate of change in NIHSS during hospitalization, by treatment, overall and by laterality, NUVISTA**

| **By treatment group^c^** | | **Change in NIHSS per day (95% CI)** | | | |
| --- | --- | --- | --- | --- | --- |
|  |  | **Overall** | | **By laterality** | |
|  |  | **Unadjusted****^a^** | **Adjusted^a,b^** | **Left laterality, Adjusted^a,b^** | **Right laterality,**  **Adjusted^a,b^** |
|  | **Treatment** | -1.059 (-1.510, -0.609) | -1.038 (-1.489, -0.588) | -1.593 (-2.243, -0.943) | -0.436 (-1.020, 0.148) |
|  | **Sham** | -1.066 (-1.473, -0.660) | -1.070 (-1.477, -0.663) | -0.954 (-1.542, -0.365) | -1.218 (-1.743, -0.693) |
|  | **Difference** | 0.007 (-0.600, 0.614) | 0.031 (-0.576, 0.639) | -0.639 (-1.516, 0.237) | 0.782 (-0.003, 1.567) |
|  | **p-value^d^** | 0.98 | 0.92 | 0.15 | 0.051 |
|  |  |  |  | p_interaction_ = 0.02^f^ | |
| **By assigned treatment^e^** | |  |  |  |  |
|  | **Treatment** | -1.271 (-1.840, -0.701) | -1.250 (-1.819, -0.681) | -1.710 (-2.500, -0.919) | -0.757 (-1.540, 0.026) |
|  | **Sham** | -1.104 (-1.502, -0.707) | -1.107 (-1.503, -0.711) | -0.959 (-1.531, -0.387) | -1.275 (-1.789, -0.761) |
|  | **Difference** | -0.166 (-0.860, 0.528) | -0.143 (-0.836, 0.551) | -0.751 (-1.726, 0.224) | 0.518 (-0.417, 1.453) |
|  | **p-value^d^** | 0.64 | 0.69 | 0.13 | 0.28 |
|  |  |  |  | p_interaction_ = 0.07^f^ | |
| **NIHSS through day 5, by assigned treatment^f^** | |  |  |  |  |
|  | **Treatment** | -1.624 (-2.586, -0.662) | -1.613 (-2.576, -0.651) | -2.204 (-3.212, -1.196) | -0.501 (-1.496, 0.493) |
|  | **Sham** | -1.568 (-2.154, -0.983) | -1.562 (-5.25, -0.978) | -1.185 (-1.934, -0.436) | -1.525 (-2.071, -0.979) |
|  | **Difference** | -0.056 (-1.181, 1.069) | -0.052 (-1.176, 1.072) | -1.019 (-2.275, 0.237) | 1.024 (-0.109, 2.156) |
|  | **p-value^d^** | 0.92 | 0.93 | 0.11 | 0.08 |
|  |  |  |  | p_interaction_ = 0.02^f^ | |

^a^ Estimated using a mixed model that includes a random effect for person, as the rate of change in NIHSS (change per day) during hospitalization, i.e., until discharge from the hospital or transition to hospice (limited through day 5 only when noted). A negative rate of change indicates improvement on average on the NIHSS score over this time period.

^b^ Adjusted for percent reperfusion and time between last known normal and attempted recanalization.

^c^ Based on all NIHSS scores, except day 30 NIHSS (N=215): 100 NIHSS scores from 17 participants in the treated group and 115 NIHSS scores from 18 participants in the sham group, including 11 in the treated group with left laterality (60 NIHSS scores), 6 in the treated group with right laterality (40 NIHSS scores), 8 in the sham group with left laterality (56 NIHSS scores), and 10 in the sham group with right laterality (59 NIHSS scores).

^d^ p-value for the interaction between treatment and time, which tests whether the treatment and sham groups have different rates of change in the NIHSS, which corresponds with the 95% CI for the difference estimated in the line above.

^e^ Based on all NIHSS scores, except day 30 NIHSS (N=215), as above but with treatment as a time-varying variable until randomization (79 post-randomization NIHSS scores among those in the treatment group overall and 136 NIHSS scores overall among those in the treatment group prior to randomization or among those in the sham group).

^f^ Restricted to the 199 (92.6%) NIHSS scores from the first 5 full days after LKN (the approximate period of treatment), with treatment as a time-varying variable until randomization (72 post-randomization NIHSS scores among those in the treatment group overall and 127 NIHSS scores overall among those in the treatment group prior to randomization or among those in the sham group).

^g^ p-value for 3-way interaction, between laterality and the treatment-time (rate of change) interaction, obtained from a single mixed effects model and person as a random effect, and which tests whether the association between treatment and time (rate of change) depends on laterality.

Abbreviations: CI = confidence interval; LKN = last known normal; NIHSS = National Institutes of Health stroke scale.

**Supplemental Table 3.** **Cytokine levels in relation to time from last known normal, by treatment, overall and by laterality**

| **Cytokine** | **Treatment** | **Unit of time from LKN** | **Adjusted^a^ difference in cytokine per specified time unit (95% CI)** | | |
| --- | --- | --- | --- | --- | --- |
|  |  |  | **All strokes^b^** | **By laterality^c^** | |
|  |  |  |  | **Left laterality** | **Right laterality** |
| **IL1β** |  |  |  |  |  |
|  | **Treatment** | Days | -0.222 (-0.903 ,0.460) | -0.218 (-0.837, 0.400) | -0.346 (-1.821, 1.128) |
|  |  | Days squared | 0.030 (-0.057, 0.116) | 0.031 (-0.048, 0.111) | 0.058 (-0.126, 0.242) |
|  | **Sham** | Days | 0.327 (-0.047, 0.702) | 0.142 (-0.258, 0.542) | 0.490 (-0.146, 1.126) |
|  |  | Days squared | -0.055 (-0.111, 0.002) | -0.038 (-0.098, 0.021) | -0.061 (-0.158, 0.035) |
|  | **Interaction p-value****^d^** |  | 0.24 | 0.17 | 0.52 |
| **IL6** |  |  |  |  |  |
|  | **Treatment** | Days | -4.099 (-7.655, -0.543) | -5.006 (-9.519, -0.493) | -2.322 (-7.897, 3.253) |
|  |  | Days squared | 0.372 (-0.078, 0.823) | 0.477 (-0.099, 1.054) | 0.150 (-0.548, 0.848) |
|  | **Sham** | Days | 4.323 (2.369, 6.277) | 5.722 (2.781, 8.663) | 2.658 (0.188, 5.129) |
|  |  | Days squared | -0.585 (-0.878, -0.292) | -0.774 (-1.214, -0.335) | -0.355 (-0.729, 0.020) |
|  | **Interaction p-value^d^** |  | **0.0001** | **0.0005** | 0.14 |
| **IL10** |  |  |  |  |  |
|  | **Treatment** | Days | -0.078 (-0.412, 0.255) | -0.159 (-0.499, 0.182) | 0.005 (-0.651, 0.661) |
|  |  | Days squared | 0.014 (-0.028, 0.057) | 0.029 (-0.015, 0.072) | 0.004 (-0.078, 0.086) |
|  | **Sham** | Days | 0.150 (-0.034, 0.333) | 0.101 (-0.119, 0.320) | 0.206 (-0.077, 0.488) |
|  | **Interaction p-value^d^** |  | 0.25 | 0.07 | 0.83 |
| **IL17α** |  |  |  |  |  |
|  | **Treatment** | Days | -0.158 (-1.864, 1.547) | -0.023 (-1.650, 1.604) | -0.711 (-4.306, 2.885) |
|  |  | Days squared | 0.041 (-0.175, 0.257) | 0.039 (-0.169, 0.248) | 0.117 (-0.332, 0.566) |
|  | **Sham** | Days | 0.432 (-0.506, 1.370) | 0.139 (-0.914, 1.191) | 0.616 (-0.949, 2.182) |
|  |  | Days squared | -0.065 (-0.206, 0.076) | -0.038 (-0.194, 0.119) | -0.066 (-0.303, 0.172) |
|  | **Interaction p-value^d^** |  | 0.57 | 0.31 | 0.78 |
| **TNF-α** |  |  |  |  |  |
|  | **Treatment** | Days | -0.120 (-0.597, 0.357) | 0.028 (-0.450, 0.507) | -0.427 (-1.400, 0.546) |
|  |  | Days squared | 0.018 (-0.043, 0.078) | -0.001 (-0.062, 0.060) | 0.062 (-0.060, 0.184) |
|  | **Sham** | Days | 0.225 (-0.038, 0.488) | 0.148 (-0.163, 0.458) | 0.271 (-0.152, 0.695) |
|  |  | Days squared | -0.037 (-0.077, 0.002) | -0.033 (-0.079, 0.013) | -0.035 (-0.100, 0.029) |
|  | **Interaction p-value^d^** |  | 0.26 | 0.30 | 0.38 |
| **Cytokine z-score^e^** |  |  |  |  |  |
|  | **Treatment** | Days | -0.240 (-0.661, 0.181) | -0.297 (-0.678, 0.084) | -0.243 (-1.142, 0.657) |
|  |  | Days squared | 0.030 (-0.024, 0.083) | 0.038 (-0.011, 0.087) | 0.033 (-0.079, 0.146) |
|  | **Sham** | Days | 0.304 (0.072, 0.536) | 0.293 (0.047, 0.540) | 0.305 (-0.090, 0.700) |
|  |  | Days squared | -0.046 (-0.080, -0.011) | -0.049 (-0.086, -0.013) | -0.038 (-0.098, 0.022) |
|  | **Interaction p-value^d^** |  | 0.07 | **0.02** | 0.55 |
| **Cytokine z-score w/o IL10** |  |  |  |  |  |
|  | **Treatment** | Days | -0.259 (-0.659, 0.140) | -0.285 (-0.653, 0.084) | -0.292 (-1.137, 0.554) |
|  |  | Days squared | 0.029 (-0.021, 0.080) | 0.033 (-0.014, 0.080) | 0.037 (-0.069, 0.143) |
|  | **Sham** | Days | 0.323 (0.103, 0.544) | 0.322 (0.083, 0.560) | 0.300 (-0.074, 0.674) |
|  |  | Days squared | -0.049 (-0.082, -0.016) | -0.053 (-0.088, -0.018) | -0.038 (-0.095, 0.018) |
|  | **Interaction p-value^d^** |  | **0.04** | **0.03** | 0.46 |

^a^ Adjusted for time from LKN to recanalization (or similar reference time), percent reperfusion, and baseline NIHSS as fixed effects, in addition to plate and patient as random effects in a mixed model with the specified cytokine as the outcome; treatment and time from LKN and their product term as fixed effects; and laboratory plate and patient as random effects.

^b^ Based on 132 measurements for each cytokine: 63 cytokine measurements from 17 patients in the treatment group (14 before randomization and 49 after randomization) and 69 measurements from 18 patients in the sham group, with treatment as a time-varying variable prior to randomization and per assignment after randomization.

^c^ Based on 72 measurements for each cytokine (39 measurements from the treatment group and 33 from the sham group) from patients with left sided stroke.

^d^ Based on 60 measurements for each cytokine (24 measurements from the treatment group and 36 from the sham group) from patients with right sided stroke.

^e^ Mean of the z-score for each of the five cytokines, as an overall summary measure.

Abbreviations: CI = confidence interval; IL = interleukin; LKN = last known normal (prior to stroke); Health Stroke Scale; TNF = tumor necrosis factor

**Supplementary Material References**

1.	Pfister IB, Zandi S, Gerhardt C, Spindler J, Reichen N, Garweg JG. Risks and Challenges in Interpreting Simultaneous Analyses of Multiple Cytokines. *Translational vision science & technology*. 2020;9:27. doi: 10.1167/tvst.9.7.27

2.	Garcia TP, Marder K. Statistical Approaches to Longitudinal Data Analysis in Neurodegenerative Diseases: Huntington's Disease as a Model. *Curr Neurol Neurosci Rep*. 2017;17:14. doi: 10.1007/s11910-017-0723-4

3.	Audebert HJ, Rott MM, Eck T, Haberl RL. Systemic Inflammatory Response Depends on Initial Stroke Severity but Is Attenuated by Successful Thrombolysis. *Stroke*. 2004;35:2128-2133. doi: 10.1161/01.STR.0000137607.61697.77

4.	U.S. Food and Drug Administration. Adjusting for Covariates in Randomized Clinical Trials for Drugs and Biological Products Guidance for Industry. 2023

5.	LeCouffe NE, Kappelhof M, Treurniet KM, Lingsma HF, Zhang G, van den Wijngaard IR, van Es ACGM, Emmer BJ, Majoie CBLM, Roos YBWEM, et al. 2B, 2C, or 3. *Stroke*. 2020;51:1790-1796. doi: 10.1161/STROKEAHA.119.028891

6.	Huguenard AL, Tan G, Rivet DJ, Gao F, Johnson GW, Adamek M, Coxon AT, Kummer TT, Osbun JW, Vellimana AK, et al. Auricular vagus nerve stimulation for mitigation of inflammation and vasospasm in subarachnoid hemorrhage: a single-institution randomized controlled trial. *J Neurosurg*. 2025:1-12. doi: 10.3171/2024.10.Jns241643
